# Connectomics-guided meta-learning for decoding and anticipatory prediction of sleep spindles from basal ganglia local field potentials in Parkinson’s disease

**DOI:** 10.64898/2026.04.01.26349783

**Authors:** Chenfei Ye, Jiahui Liao, Zixiao Yin, Yue Li, Yichen Xu, Houyou Fan, Ting Ma, Jianguo Zhang

## Abstract

Sleep disturbances are pervasive, debilitating non-motor symptoms of Parkinson’s disease (PD), where sleep spindle deficits directly drive cognitive decline and disease progression. Current adaptive deep brain stimulation (aDBS) for PD is largely limited to motor symptom management, with no established technical foundation for sleep spindle-targeted closed-loop modulation. The functional role of the basal ganglia in human sleep spindle regulation remains incompletely characterized, and no robust cross-subject pipeline exists to decode these transient events from clinically implanted DBS electrodes. Here, we developed a connectomics-guided meta-learning framework for cross-subject sleep spindle decoding and anticipatory prediction, using whole-night synchronized basal ganglia local field potential and polysomnography data from 17 PD patients with bilateral DBS implants. Our framework achieved 92.63% accuracy for concurrent spindle decoding and 83.44% accuracy for 2-second-ahead prediction, with optimal signals localized to the limbic subthalamic nucleus and <50 ms total latency meeting real-time closed-loop requirements. This work defines the neuroanatomical substrate of basal ganglia spindle signaling in PD, establishes the cross-subject spindle decoding pipeline for clinical DBS systems, and provides a critical translational foundation for sleep-targeted closed-loop aDBS to mitigate PD non-motor burden.

## Introduction

Sleep disturbances are a pervasive and debilitating non-motor symptom in Parkinson’s disease (PD), over 80% of patients and significantly accelerating cognitive decline (1–3). While pathological beta oscillations (13–30 Hz) in the awake state are well-characterized biomarkers of motor impairment (4–7), the sleeping PD brain exhibits distinct electrophysiological disruptions. Recent high-impact studies have revealed that excessive pallidal beta activity persists during sleep and is directly correlated with sleep fragmentation and insomnia severity (8, 9). However, beyond these broad spectral abnormalities, finer-grained non-rapid eye movement (NREM) sleep micro-events—specifically sleep spindles (11–16 Hz thalamocortical bursts)—are critical for memory consolidation and synaptic plasticity (10–12). In PD, sleep spindles are often blunted, with reduced density and altered morphology, which correlates with impaired off-line learning and cognitive deficits(13). Crucially, emerging evidence from human closed-loop stimulation studies demonstrates that precisely synchronizing stimulation with endogenous sleep rhythms (e.g., slow waves or spindles) can enhance memory consolidation and restore physiological sleep architecture(14). This suggests that spindles are not merely passive markers but actionable therapeutic targets for next-generation therapies.

While polysomnography (PSG) remains the gold standard for capturing these events, its complexity limits its use to clinical snapshots rather than longitudinal monitoring. Deep Brain Stimulation (DBS) offers a unique “window” into the brain, allowing for chronic sensing of intracranial Local Field Potentials (LFPs) directly from the basal ganglia(9). Recent findings indicate that the basal ganglia (BG) are not just motor relays but actively participate in sleep regulation. For instance, *Zhang et al.* recently identified that pathological beta bursts in the subthalamic nucleus (STN) competitively interfere with physiological sleep spindles, providing a causal mechanistic link between basal ganglia pathophysiology and sleep disruption in PD (15). Despite these advances, current adaptive DBS (aDBS) research and clinical translation remain overwhelmingly focused on beta-based feedback to treat awake motor symptoms (16, 17). While pioneering efforts have extended aDBS to sleep, notably work by *Smyth et al.* demonstrating the feasibility of an aDBS algorithm that modulates stimulation parameters in response to intracranially classified sleep stages (18), these macroscopic approaches lack the temporal resolution to target transient, millisecond-scale sleep spindle events. The critical absence of robust, LFP-based spindle detection and prediction methods has thus far precluded the development of precision neuroprosthetics capable of both restoring spindle-mediated plasticity and alleviating sleep disturbances in PD.

To address these unmet clinical and technical challenges, we leveraged a unique, synchronized whole-night basal ganglia LFP-PSG dataset from 17 PD patients with bilateral DBS implants targeting either the STN or globus pallidus internus (GPi). First, we developed a functional connectomics-guided framework for automated optimal LFP channel selection. The core motivation underpinning this design is to address a key clinical barrier of conventional channel screening approaches, which rely on laborious, time-consuming full-channel pre-testing incompatible with routine clinical workflow. This framework enables individualized, data-driven identification of the most informative recording channel for each patient without extensive pre-testing. Building on this standardized channel selection pipeline, we designed a meta-learning convolutional neural network (MetaCNN) to mitigate the profound inter-subject variability in LFP signal characteristics that has long limited the cross-subject generalizability and clinical translation of conventional LFP decoding models. This meta-learning paradigm optimizes for the extraction of generalizable, subject-invariant features of sleep spindles, enabling rapid, robust adaptation to unseen patients using only a minimal calibration dataset. The resulting integrated pipeline not only enables robust cross-subject decoding of NREM sleep spindles from basal ganglia LFP signals, but also supports reliable anticipatory prediction of upcoming spindle events prior to their cortical manifestation. Our work not only establishes the first high-performance framework for decoding and predicting sleep spindles from the human basal ganglia, but also delineates the critical electrophysiological and neuroanatomical substrates of sleep spindle regulation in the parkinsonian brain, laying essential preclinical groundwork for next-generation closed-loop sleep neuromodulation therapies targeting the debilitating non-motor burden of PD.

## Results

### Participant Characteristics and Clinical Profiles

Seventeen patients with idiopathic PD were enrolled in this study, including 8 males and 9 females with a mean age of 57.2 ± 12.0 years and a mean disease duration of 9.9 ± 5.2 years. Thirteen patients underwent bilateral DBS implantation targeting the STN, while the remaining 4 were implanted at the GPi (detailed demographic and clinical characteristics are summarized in Tables 1 and 2).

**Table 1.** Demographics and clinical characteristics of the included patients (Individual-level)

| Patient | Target | Gender | Age ranges (years) | DD (years) | PSQI | UPDRS-III (on) | UPDRS-III (off) |
| --- | --- | --- | --- | --- | --- | --- | --- |
| Sub-01 | STN | Female | 51-55 | 23 | 6 | 15 | 22 |
| Sub-02 | STN | Female | 61-65 | 8 | 15 | 30 | 37 |
| Sub-03 | STN | Male | 66-70 | 8 | 9 | 16 | 34 |
| Sub-04 | STN | Male | 66-70 | 4 | 12 | 25 | 51 |
| Sub-05 | STN | Male | 36-40 | 20 | 13 | 34 | 54 |
| Sub-06 | STN | Male | 71-75 | 7 | 8 | 4 | 24 |
| Sub-07 | STN | Female | 51-55 | 6 | 7 | 25 | 39 |
| Sub-08 | STN | Female | 71-75 | 11 | 3 | 18 | 30 |
| Sub-09 | STN | Male | 36-40 | 13 | 16 | 36 | 74 |
| Sub-10 | STN | Male | 56-60 | 6 | 9 | 38 | 69 |
| Sub-11 | STN | Female | 76-80 | 15 | 4 | 28 | 42 |
| Sub-12 | STN | Female | 61-65 | 8 | 6 | 4 | 19 |
| Sub-13 | STN | Female | 56-60 | 11 | 13 | 21 | 34 |
| Sub-14 | GPI | Female | 46-50 | 6 | 5 | 32 | 67 |
| Sub-15 | GPI | Male | 36-40 | 9 | 15 | 4 | 24 |
| Sub-16 | GPI | Female | 56-60 | 8 | 17 | 37 | 66 |
| Sub-17 | GPI | Male | 51-55 | 5 | 5 | 23 | 38 |
| Mean ± SD | / | / | 57.2 ± 12.0 | 9.9 ± 5.2 | 9.6 ± 4.6 | 22.9 ± 11.5 | 42.6 ± 17.8 |

3 *DD*: duration of disease, *PSQI*: Pittsburgh sleep quality index, *UPDRS-III*: the MDS-Unified 4 Parkinson's Disease Rating Scale-III.

**Table 2.** Demographics and clinical characteristics of the included patients (Group-level)

| Variable | All Patients<br>(n=17) | STN (n=13) | GPi (n=4) | T-test |  |
| --- | --- | --- | --- | --- | --- |
|  |  |  |  | t | p |
| Age (years) | 57.2±12.0 | 59.8±12.6 | 49.8±8.3 | 1.48 | 0.080 |
| DD (years) | 9.9±5.2 | 10.8±5.9 | 7.0±1.8 | 2.07 | 0.027 |
| PSQI | 9.6±4.6 | 9.0±4.2 | 10.5±6.4 | -0.44 | 0.331 |
| UPDRS-III<br>on | 22.9±11.5 | 22.6±11.0 | 24.0±14.5 | -0.21 | 0.420 |
| UPDRS-III<br>off | 42.6±17.8 | 41.0±17.0 | 48.8±21.3 | -0.78 | 0.223 |

3 *DD*: duration of disease, *PSQI*: Pittsburgh sleep quality index, *UPDRS-III*: the MDS-Unified 4 Parkinson's Disease Rating Scale-III.

All participants reported poor subjective sleep quality, with a mean Pittsburgh Sleep Quality Index (PSQI) score of 9.6 ± 4.6. DBS treatment yielded a significant improvement in motor symptoms: the mean Unified Parkinson’s Disease Rating Scale part III (UPDRS-III) score was 22.9 ± 11.5 with DBS switched on, compared to 42.6 ± 17.8 with DBS off, corresponding to a ∼46% reduction in motor impairment (p < 0.001, Wilcoxon signed-rank test).

Between-group comparisons revealed that the STN cohort had a significantly longer disease duration than the GPi cohort (10.8 ± 5.9 vs. 7.0 ± 1.8 years; t = 2.07, p = 0.027, independent samples t-test). No significant between-group differences were observed for age (59.8 ± 12.6 vs. 49.8 ± 8.3 years; t = 1.48, p = 0.080), subjective sleep quality (PSQI: 9.0 ± 4.2 vs. 10.5 ± 6.4; t = –0.44, p = 0.331), or motor symptom severity (UPDRS-III with DBS on: 22.6 ± 11.0 vs. 24.0 ± 14.5, t = –0.21, p = 0.420; UPDRS-III with DBS off: 41.0 ± 17.0 vs. 48.8 ± 21.3, t = –0.78, p = 0.223). These findings support the pooled analysis of both cohorts where specified in subsequent analyses.

### Sleep Spindle Identification via Simultaneous EEG-LFP Recordings

To establish ground-truth labels for subsequent spindle decoding models, we performed sleep staging and automated sleep spindle detection using overnight simultaneous polysomnography (PSG) and basal ganglia local field potential (LFP) recordings (full preprocessing and detection protocols are detailed in the Methods section).

Manual sleep staging yielded 4–6 hours of artifact-free non-rapid eye movement (NREM; stages N2 and N3) data per participant. Automated spindle detection via the YASA toolbox identified 108 to 424 valid spindle events per subject, 97.4% of which occurred during N2 sleep, with the remaining 2.6% during N3 sleep (Supplementary Table 1). Spatial-spectral characterization revealed that 72.0% of detected spindle events involved frontal derivations (F3/F4), with a mean peak frequency of 12.91 Hz consistent with the canonical profile of slow sleep spindles (11–13 Hz). All validated events exhibited prominent sigma-band (11–16 Hz) activity, with a mean RMS amplitude 2.6-fold higher than baseline, confirming the fidelity of our detection pipeline. Based on these results, we extracted corresponding 2-second LFP segments to generate a balanced dataset of ‘spindle-positive’ and ‘non-spindle’ labels for downstream decoding analyses.

### Connectomics-guided Framework Identifies Optimal LFP Channels

With the ground-truth spindle labels and paired LFP dataset established, we first developed a functional connectomics-guided framework for automated optimal LFP channel selection as the foundational step of our cross-subject sleep spindle decoding pipeline (Figure 1d). This framework was designed to address the key clinical limitation of conventional channel screening approaches, which rely on laborious, time-consuming full-channel pre-testing that is incompatible with routine clinical workflow. It was implemented within a strict leave-one-subject-out (LOSO) cross-validation pipeline, with full protocol details provided in the Methods section and Supplementary Figure 1. At the core of this framework, we defined a group-level representation map (R-map): a statistical whole-brain functional connectivity atlas derived from the training cohort, which delineates the whole-brain network profile associated with high-fidelity sleep spindle signal capture from basal ganglia LFP signals.

**Figure 1.**
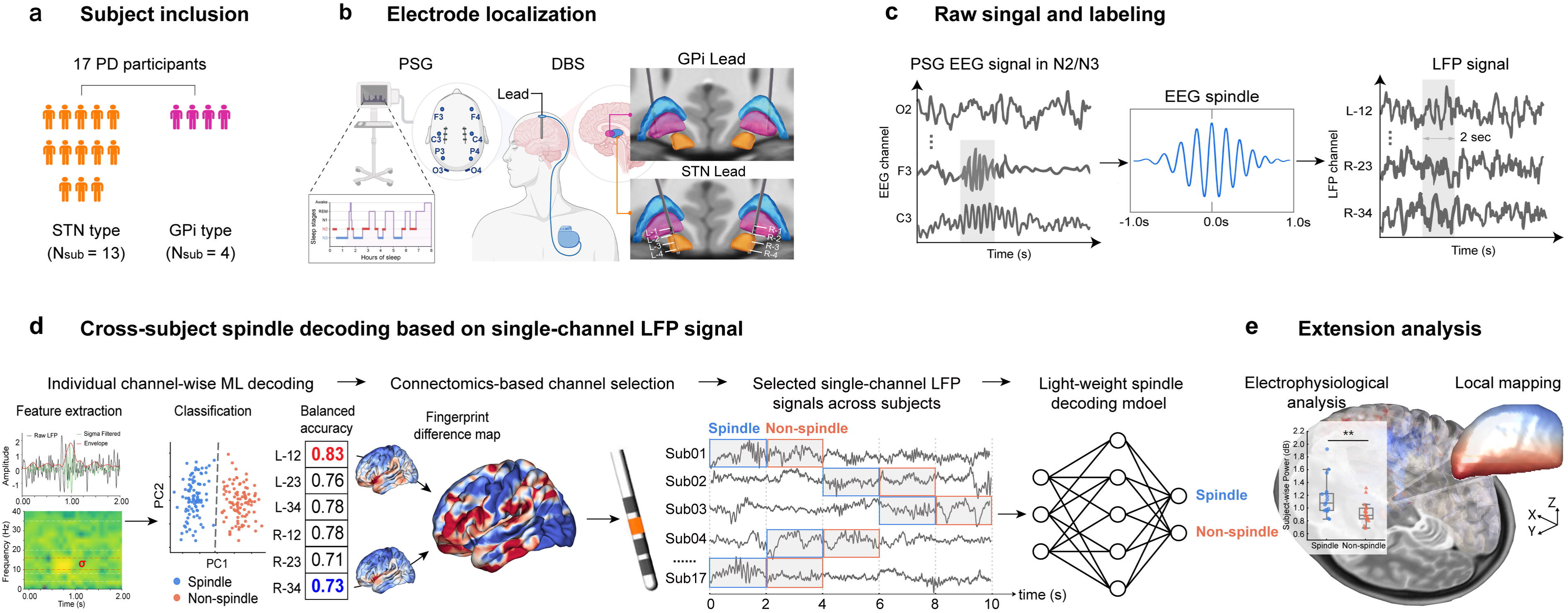
Overview of the connectomics-guided meta-learning framework for cross-subject sleep spindle decoding from basal ganglia local field potentials. **a.** Cohort profile including 17 participants with idiopathic Parkinson’s disease (PD), 13 with deep brain stimulation (DBS) implants targeting the subthalamic nucleus (STN) and 4 targeting the globus pallidus internus (GPi). **b.** Experimental setup for simultaneous whole-night polysomnography (PSG) and bilateral basal ganglia local field potential (LFP) recordings. **c.** Ground-truth sleep spindle labeling paradigm: spindle events were algorithmically identified and validated from scalp electroencephalography (EEG) during non-rapid eye movement (NREM) sleep stages N2 and N3, with paired LFP segments extracted for model training and validation. **d.** The proposed cross-subject decoding pipeline, integrating a functional connectomics-guided framework for patient-specific optimal LFP channel selection, followed by a lightweight meta-learning convolutional neural network (MetaCNN) for robust spindle detection and prediction. **e.** Downstream validation analyses, including electrophysiological feature characterization and spatial mapping of decoding performance across the basal ganglia.

Whole-cohort consensus R-map analysis revealed a highly polarized network topology underlying decoding efficacy, with statistically significant peak T-statistics (Tmax = 4.46, Tmin = -4.08, both p < 0.001; Supplementary Figure 2). Higher spindle decoding performance was significantly positively associated with functional connectivity to prefrontal and limbic hubs, most prominently the orbitofrontal cortex and anterior cingulate cortex. Conversely, lower decoding performance was significantly associated with stronger functional connectivity to primary sensorimotor areas and the superior parietal cortex.

Quantitative validation confirmed the robustness of our connectomics-guided channel matching process: channels with the highest R-map match had significantly higher spatial correlations with the training-derived group-level R-map (median r = 0.149, interquartile range [IQR]: 0.116–0.183) than channels with the lowest R-map match (median r = 0.065, IQR: 0.018–0.101; subject-wise correlation coefficients are provided in Supplementary Table 2). The channel exhibiting the highest spatial correlation to the R-map was automatically selected as the optimal channel for each test subject, and was used exclusively for all downstream decoding analyses.

### Meta-Learning Enhances Cross-Subject Decoding and Real-Time Execution

Having identified the optimal single LFP channel for each participant via our connectomics-guided framework, we next developed a meta-learning convolutional neural network (MetaCNN) to decode sleep spindle events directly from these high-fidelity, biologically targeted single-channel signals. While our channel selection pipeline mitigates inter-contact signal variability and enriches spindle-related neural signatures in the model input, conventional deep learning models remain limited by profound inter-subject variability in LFP signal characteristics, which restricts their cross-subject generalizability and real-world clinical utility. Our MetaCNN addresses this core barrier via the model-agnostic meta-learning (MAML) paradigm, which optimizes for a highly generalizable, easily adaptable parameter initialization, enabling rapid and robust adaptation to unseen patients using only a minimal, few-shot calibration dataset (support set).

Evaluated using LOSO cross-validation, the MetaCNN achieved excellent cross-subject spindle classification performance, with a mean accuracy of 92.63 ± 3.13%, F1-score of 92.19 ± 3.32%, sensitivity of 89.79 ± 4.61%, and specificity of 95.47 ± 4.24% (Figure 2b, Supplementary Table 4). These performance metrics were significantly higher than those achieved by two benchmark models: a standard convolutional neural network without meta-learning (BaseCNN; accuracy: 76.22 ± 6.67%, F1-score: 75.65 ± 5.95%, sensitivity: 73.18 ± 5.31%, specificity: 79.27 ± 11.69%, p < 0.01, Supplementary Table 5), and a feature-based support vector machine (SVM) using spectral power and phase-locking value (PLV) as inputs (accuracy: 71.10 ± 6.91%, F1-score: 68.70 ± 11.30%, sensitivity: 68.50 ± 22.77%, specificity: 73.71 ± 23.18%, p < 0.01, Supplementary Table 6).

**Figure 2.**
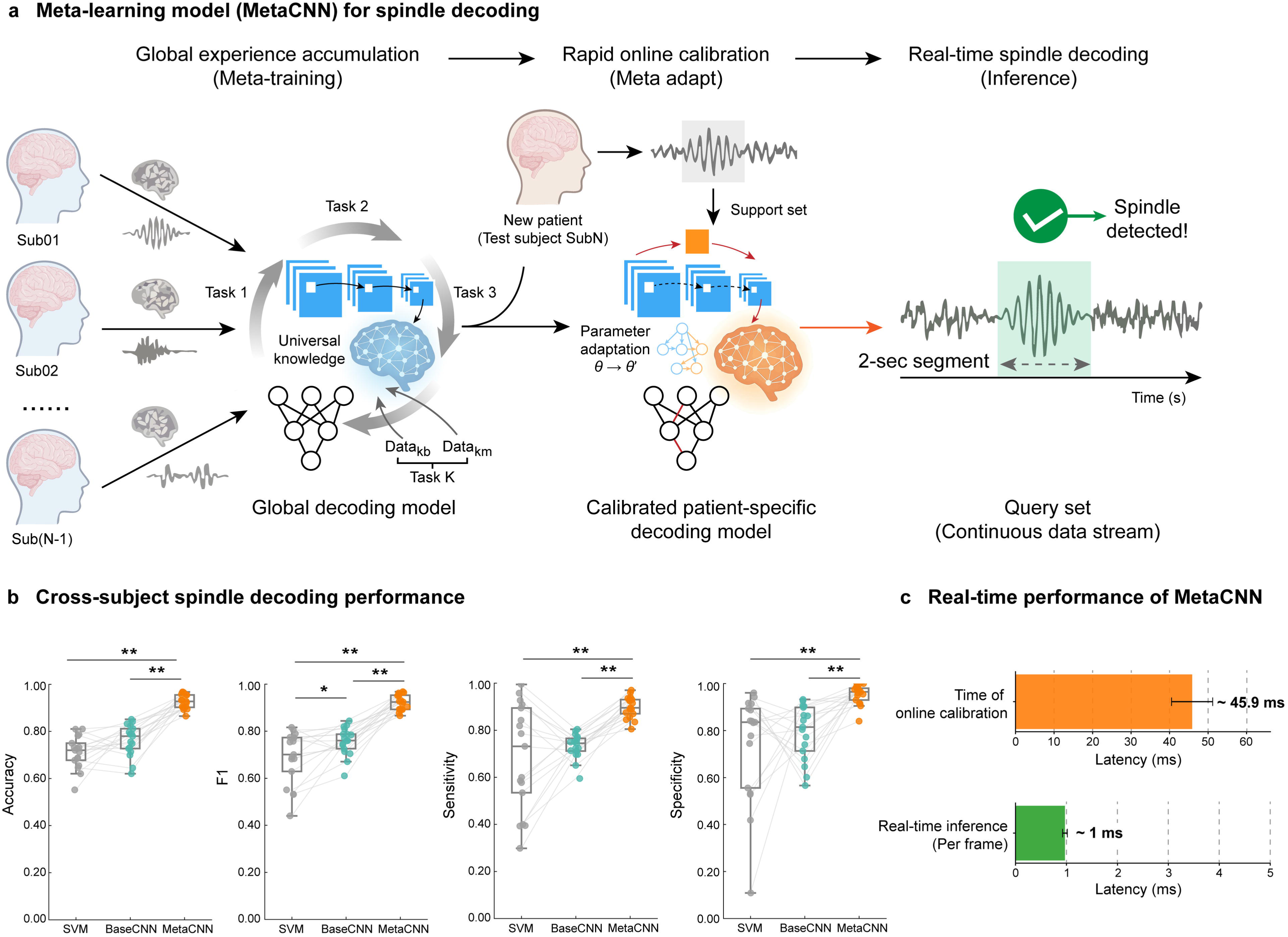
Meta-learning framework enables superior cross-subject decoding performance and real-time clinical feasibility. **a.** Schematic of the MetaCNN architecture, including a meta-training phase to learn generalizable, subject-invariant spindle representations, an online calibration phase (meta-adaptation) using a minimal support set from an unseen patient, and continuous real-time inference. **b**. Cross-subject decoding performance comparisons between the support vector machine (SVM), baseline convolutional neural network (BaseCNN), and MetaCNN, quantified by accuracy, F1-score, sensitivity, and specificity (\**p* < 0.05, \*\**p* < 0.01; paired t-test/Wilcoxon signed-rank test). Gray lines represent the performance trajectories of individual subjects. **c.** Computational latency evaluation of the MetaCNN, including the time required for online meta-adaptation and single-frame real-time inference. For all box plots, center lines indicate the median, box bounds represent the 25th and 75th percentiles, and whiskers extend to 1.5× the interquartile range (IQR).

Beyond decoding performance, the clinical feasibility of closed-loop neuromodulation is critically dependent on algorithmic latency and computational footprint. Notably, the MetaCNN has an ultra-lightweight architecture, with only ∼3,500 trainable parameters (0.0035M). Computational efficiency analyses demonstrated that the rapid online calibration phase (meta-adaptation), using the minimal few-shot support set from a new subject, required only 45.86 ± 2.47 ms to complete (Figure 2c, Supplementary Table 7, Supplementary Figure 3). Furthermore, the real-time inference latency for processing a single 2-second query frame was only 0.97 ± 0.42 ms. This ultra-low latency confirms that the MetaCNN is a viable computational engine for real-time adaptive DBS systems.

### Optimal Channel Exhibit Distinct Limbic Proximity and Electrophysiological Enhancements

We identified distinct electrophysiological and anatomical signatures of the connectomics-guided optimal LFP channels, which underpin their superior sleep spindle decoding performance (Figure 3).

**Figure 3.**
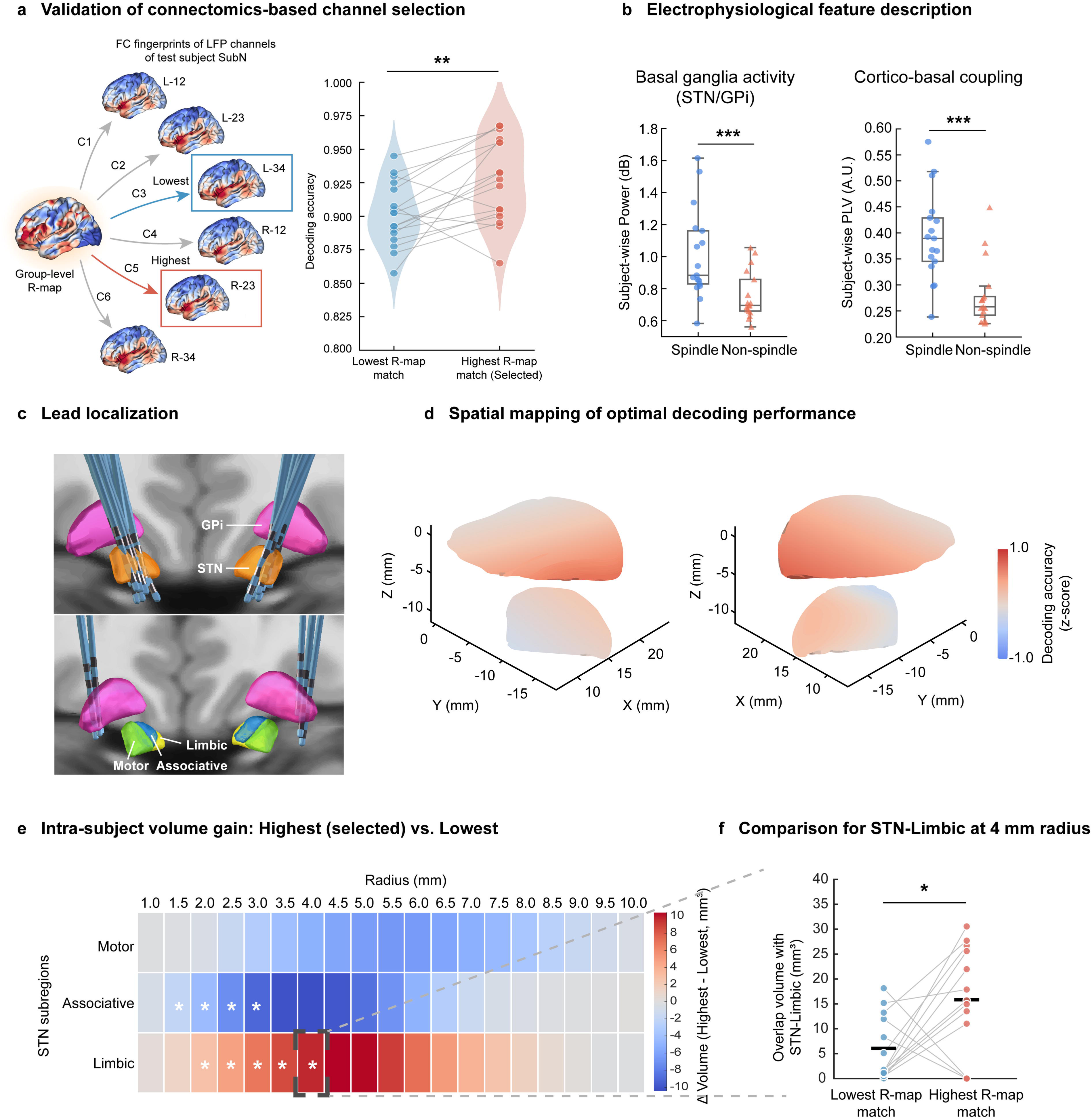
Connectomics-guided optimal channels show distinct limbic network proximity and enhanced spindle-related electrophysiological signatures. **a.** Comparison of MetaCNN decoding accuracy between channels with the highest and lowest representation map (R-map) matches (\**p* < 0.05; paired t-test / Wilcoxon signed-rank test). **b**. Comparisons of spindle-related electrophysiological features, including sigma-band peak power and cortico-basal phase-locking value (PLV), between spindle and non-spindle LFP segments (\*\**p* < 0.001; paired t-test / Wilcoxon signed-rank test). **c.** 3D reconstruction of DBS leads within the STN and GPi, with anatomical parcellation of STN subregions (motor, associative, limbic). **d.** Spatial mapping of selected single-channel decoding performance (Z-scored) across the basal ganglia geometry. **e.** Heatmap of within-subject differences in overlap volume (ΔVolume) between highest and lowest R-map matched channels across STN subregions at varying spherical volume of tissue activated (VTA) radii. **f.** Comparison of VTA overlap volume with the limbic STN subregion at a standardized 4-mm radius (\**p* < 0.05; two-tailed paired t-test / Wilcoxon signed-rank test). For all box plots, center lines indicate the median, box bounds represent the 25th and 75th percentiles, and whiskers extend to 1.5× the IQR.

First, we validated the functional superiority of our connectomics-guided channel selection strategy. Within the same test subjects, MetaCNN achieved significantly higher decoding accuracy using channels with the highest R-map match, compared to channels with the lowest R-map match (P = 0.005; Figure 3a, Supplementary Tables 4 and 8).

Second, optimal channels exhibited robust spindle-related electrophysiological signatures at both local and network levels. During spindle events, LFP signals from optimal channels showed a significant elevation in sigma-band (11–16 Hz) periodic peak power, compared to non-spindle baseline segments (subject-wise mean: 1.009 ± 0.281 vs. 0.756 ± 0.150 dB; p = 6.107 × 10□□; Figure 3b, Supplementary Table 9). Concurrently, cortico-basal phase synchronization (quantified via phase-locking value, PLV) between LFP and scalp EEG was significantly higher during spindle events (subject-wise mean: 0.393 ± 0.086 vs. 0.280 ± 0.062; p = 1.820 × 10□□; Figure 3b, Supplementary Table 9).

Third, high-resolution anatomical mapping revealed a clear subregional preference of high-performance channels within the STN. Spatial mapping of z-scored decoding accuracy identified robust efficacy hotspots clustered in the ventromedial STN. Subregional analysis (detailed in Methods) showed that channels with the highest R-map match had significantly greater volume overlap with the limbic subregion of the STN, compared to lowest-match channels (p < 0.05 at 4-mm VTA radius; Figure 3f, Supplementary Table 10). In contrast, optimal channels showed significantly less overlap with the associative STN subregion (negative ΔVolume, p < 0.05 at 2.0–3.0 mm radii; Supplementary Table 10), reflecting clear spatial segregation along the DBS lead trajectory.

Collectively, these findings demonstrate that our connectomics-guided framework reliably identifies channels positioned at the limbic-prefrontal circuit interface of the basal ganglia, which carry robust spindle-related neural signatures to support high-accuracy decoding.

### Meta-Learning Enables Cross-Subject Decoding of Upcoming Sleep Spindles

Building on its strong performance in concurrent sleep spindle classification, our MetaCNN framework further enables reliable cross-subject anticipatory prediction of upcoming cortical sleep spindles using basal ganglia LFP signals — a critical capability that provides the temporal window required for proactive closed-loop DBS neuromodulation in next-generation adaptive systems. We evaluated the predictive performance of the unmodified MetaCNN architecture via a temporal horizon analysis, with full methodological details provided in the Methods section.

At the immediate pre-spindle horizon (Δt = 0.0 s, the 2-second window immediately preceding spindle onset; Figure 4b), MetaCNN achieved strong predictive performance with a mean accuracy of 83.44 ± 3.92%, F1-score of 83.30 ± 3.13%, sensitivity of 83.42 ± 5.44%, and specificity of 83.47 ± 5.83% (Supplementary Table 4). This performance was significantly superior to both benchmark models: the standard BaseCNN without meta-learning (accuracy: 50.66 ± 3.09%, F1-score: 50.93 ± 3.24%, P < 0.01, Supplementary Table 5) and the feature-based SVM (accuracy: 51.98 ± 5.07%, F1-score: 60.74 ± 3.90%, P < 0.01, Supplementary Table 6).

**Figure 4.**
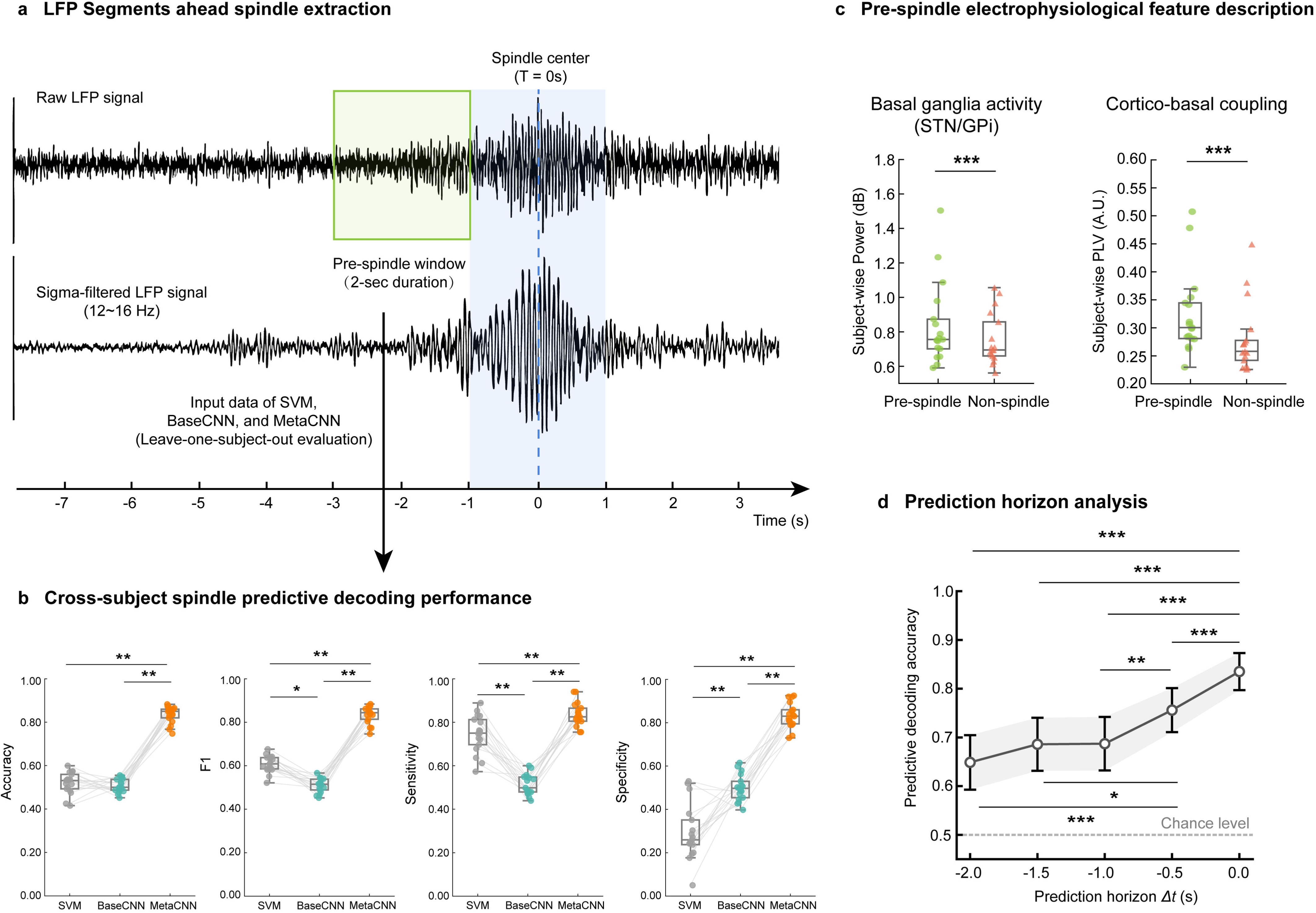
Meta-learning enables robust cross-subject anticipatory prediction of upcoming sleep spindles. **a.** Schematic of the pre-spindle predictive decoding paradigm: a 2-second LFP observation window was shifted backward relative to cortical spindle onset, with the prediction horizon (Δt) defined as the interval between the end of the observation window and spindle onset (e.g., Δt = 0.0 s indicates the window immediately preceding spindle onset). **b**. Cross-subject predictive decoding performance comparison at Δt = 0.0 s between the SVM, BaseCNN, and MetaCNN (\**p* < 0.05, \*\**p* < 0.01; paired t-test / Wilcoxon signed-rank test). Gray lines denote paired performance values for individual subjects across models. **c.** Comparison of pre-spindle electrophysiological features (peak power and cortico-basal PLV at Δt = 0.0 s) with non-spindle LFP segments (\*\**p* < 0.01; paired t-test / Wilcoxon signed-rank test). Center lines denote the median, box limits indicate the 25th and 75th percentiles, and whiskers extend to 1.5× the interquartile range. **d.** Prediction horizon analysis evaluating MetaCNN decoding accuracy across progressively earlier observation windows (Δt from 0.0 s to −2.0 s). Error bars indicate the standard deviation across subjects; the dashed line represents the 50% chance level (\**p* < 0.05, \*\**p* < 0.01, \*\*\**p* < 0.001, paired t-test / Wilcoxon signed-rank test with Bonferroni correction).

We identified significant electrophysiological signatures in the pre-spindle precursor phase that support reliable prediction. Compared to non-spindle baseline segments, the pre-spindle window was characterized by significantly elevated local sigma-band (11–16 Hz) activity in the basal ganglia (subject-wise mean power: 0.854 ± 0.250 vs. 0.756 ± 0.150 dB; P = 3.05 × 10□□), alongside a concomitant increase in cortico-basal phase synchronization (subject-wise mean PLV: 0.310 ± 0.081 vs. 0.280 ± 0.062; P = 4.30 × 10□□; Figure 4c).

Across extended prediction horizons, MetaCNN showed a progressive decrease in decoding accuracy as the observation window was shifted further back relative to spindle onset. However, the model maintained decoding performance significantly above the 50% chance level across all tested temporal windows (Δt = 0.0 s to -2.0 s; Figure 4d, Supplementary Tables 11 and 12). These findings demonstrate that early subcortical neural signatures within the basal ganglia can reliably anticipate impending cortical sleep spindles, establishing a viable temporal window for proactive closed-loop neuromodulation.

## Discussion

In this study, we established a robust, generalizable pipeline for cross-subject decoding and anticipatory prediction of NREM sleep spindles using single-channel basal ganglia local field potential LFP signals recorded from DBS electrodes in 17 patients with Parkinson’s disease PD. Leveraging a meta-learning convolutional neural network (MetaCNN) paired with a functional connectomics-guided channel selection framework, we achieved strong cross-subject generalizability with minimal subject-specific calibration. To the best of our knowledge, this is the first successful demonstration of decoding and predicting sleep spindles using exclusively basal ganglia LFP signals. This work addresses critical unmet needs in the field of adaptive DBS (aDBS), which has long focused on motor symptom management in the awake state, and provides a translational foundation for next-generation sleep-targeted neuromodulation therapies to mitigate the debilitating non-motor burden of PD.

Our functional connectomics-guided channel selection framework resolves a longstanding challenge in LFP-based decoding: profound inter-subject variability in signal characteristics, even between adjacent DBS contacts. While structural connectivity profiles are highly homogeneous across millimeter-scale adjacent contacts, functional connectivity captures state-dependent, polysynaptic network synchrony, yielding unique functional fingerprints for each contact (19–21). Our group-level representation map (R-map) reveals a highly polarized network topology underlying spindle decoding efficacy: performance is strongly predicted by connectivity to prefrontal-limbic hubs, rather than the sensorimotor circuits typically targeted in motor-focused aDBS. The spatial segregation we observed — where optimal channels target the limbic STN, while poorly performing channels overlap with the associative STN — is fully supported by the topographical organization of the cortico-basal ganglia network. Frontal slow spindles originate in medial prefrontal and anterior cingulate cortices (22), which project exclusively to the ventromedial limbic STN via the hyperdirect pathway (23). This topographical alignment confirms that our framework does not detect non-specific volume conduction, but precisely identifies the anatomical hub of sleep rhythm propagation within the basal ganglia. This work extends the existing understanding of STN functional topography, which has historically focused on motor and cognitive subregions, by establishing the limbic STN as a critical node in sleep regulation.

Our MetaCNN framework addresses a core barrier to clinical translation of LFP-based decoding: poor cross-subject generalizability of conventional models. Most existing decoding algorithms are subject-specific, requiring extensive, patient-specific calibration data that is not feasible in routine clinical practice. Our meta-learning approach optimizes for a generalizable parameter initialization, enabling robust adaptation to unseen patients with only a minimal calibration dataset. The superior cross-subject performance of our model is particularly notable in the context of the well-documented variability in sleep spindle annotation: inter-expert agreement for manual spindle scoring is only 61 ± 6% (24), indicating that our model outperforms human expert consistency even in the challenging cross-subject setting. This demonstrates that our framework captures generalizable, biologically meaningful features of sleep spindles, rather than overfitting to subject-specific noise.

Beyond decoding performance, our model’s ultra-lightweight architecture and ultra-low latency address a critical requirement for real-time closed-loop neuromodulation. The MetaCNN has only ∼3,500 trainable parameters, two orders of magnitude smaller than widely used BCI models such as DeepConvNet (25), and comparable to the minimal configuration of EEGNet (26). For clinical aDBS applications, total online adaptation latency (TOA) — the sum of calibration time (TC) and single-frame inference time (TQ) — is the key determinant of real-world feasibility (27). Our model’s TOA of ∼47 ms falls into the <50 ms “ultra-fast” latency category, suitable for high-frequency closed-loop interactions with no perceptible delay (28). This is a critical advance over existing sleep-focused aDBS approaches, which are limited to macroscopic sleep stage modulation and lack the temporal resolution to target transient spindle events. Our model’s real-time performance establishes its viability for clinical translation into implantable DBS devices with on-board processing capabilities.

The neurophysiological features driving our model’s performance provide novel mechanistic insights into the role of the basal ganglia in sleep regulation. Decoding success was most strongly predicted by two core, biologically plausible features: the magnitude of spindle-related sigma-band power modulation in the basal ganglia, and the strength of cortico-basal PLV between STN LFPs and cortical EEG during spindle events. These findings align with and substantially extend the existing mechanistic understanding of sleep spindle regulation in the human brain. Sleep spindles are generated via the cyclic inhibitory-excitatory interaction between thalamic relay nuclei and the thalamic reticular nucleus within the canonical thalamocortical loop, and the STN receives both direct and indirect anatomical projections from these same thalamic nuclei that mediate spindle initiation and propagation (29). This anatomical connectivity provides a direct biological substrate for the spindle-related LFP signatures we observed in the STN. Further, recent intracranial studies have established that phase-locking between subcortical structures and cortical spindles is a core mechanism mediating subcortical participation in NREM oscillatory dynamics: human intracranial EEG work has demonstrated that spindle-related phase synchronization coordinates hippocampal-neocortical dialogue during sleep-dependent memory consolidation (30), while recent work in PD has linked the integrity of subcortical-cortical phase coupling to the preservation of both motor and non-motor circuit function in the parkinsonian brain (31). However, our study is the first to demonstrate that these spindle-related phase-synchronized neural signals can be reliably captured from clinically standard DBS electrodes implanted in the STN, with sufficient fidelity and cross-subject consistency to support robust generalizable decoding. This finding confirms that the basal ganglia are not passive bystanders during NREM sleep, but are functionally integrated into the thalamocortical circuitry that governs sleep spindle generation, propagation, and function.

Our finding that the limbic subregion of the STN is the optimal target for spindle decoding has important implications for DBS therapy in PD. The limbic STN receives dense projections from anterior cingulate, orbitofrontal, and ventromedial prefrontal cortices, as well as indirect modulation via amygdala and ventral striatal pathways, positioning it at the interface of limbic and thalamocortical systems (23, 32). Sleep spindles orchestrate overnight hippocampal-to-neocortical transfer of emotional and declarative memories, a process that preferentially engages limbic-prefrontal networks for emotional tagging and overnight “depotentiation” of affective load while preserving factual content (33–35). Recent evidence further demonstrates spindle-mediated dialogue between hippocampus/amygdala and ventromedial prefrontal cortex (vmPFC) that selectively decouples emotional tone from memory traces during NREM sleep (30, 36). Thus, stronger spindle-related LFP signatures in this subregion likely reflect its integration of affective and memory-related signals that are preferentially synchronized with cortical spindles. The associative (central) STN subregion, which bridges basal ganglia with dorsolateral prefrontal cortex (DLPFC) to support executive control, rule-based learning, and working memory (37), exhibited moderate spindle correlations. This likely reflects spindle-driven consolidation of cognitive skills acquired during wakefulness (38), yet its contribution remains secondary to the limbic territory in our predictive and classificatory tasks. In contrast, the motor-dominant dorsolateral STN and the entire GPi showed markedly weaker decoding performance, especially in the predictive task, consistent with their primary involvement in motor control and less direct engagement in sleep-dependent emotional-memory processing. Collectively, these subregion-specific gradients reinforce the limbic STN as a privileged node for spindle orchestration during sleep-dependent memory transformation and emotional homeostasis. Precise targeting of limbic STN contacts in adaptive DBS paradigms may therefore optimise modulation of sleep architecture and affective symptoms in neuropsychiatric disorders, while underscoring the necessity of subregional electrode localization for interpreting LFP biomarkers of sleep and emotion.

Beyond concurrent spindle detection, our model’s ability to anticipate upcoming spindle events represents a critical advance for closed-loop sleep neuromodulation. Most existing closed-loop sleep stimulation protocols are reactive, delivering stimulation after a spindle is detected, which misses the opportunity to preserve or enhance spindles before they are disrupted by pathological parkinsonian beta bursts. Our model reliably predicts spindle events up to 2 seconds before their onset, with performance significantly above chance level across all tested horizons. We also identified significant pre-spindle electrophysiological signatures in the basal ganglia, including elevated sigma power and cortico-basal phase synchronization, which provide novel insights into the pre-activation of thalamocortical circuitry before spindle onset. This predictive capability enables proactive, rather than reactive, closed-loop stimulation, which is far more likely to preserve physiological sleep architecture and improve clinical outcomes.

The pipeline we present here has broad clinical and research applications. First, it enables chronic, longitudinal monitoring of sleep spindle dynamics in ambulatory PD patients, eliminating the need for repeated, resource-intensive overnight PSG. This could be used to track disease progression and cognitive decline in PD, given the well-established links between spindle loss and dementia in PD (12, 13). Second, our predictive framework enables proactive closed-loop DBS to enhance sleep spindles, with the potential to improve memory consolidation, reduce sleep fragmentation, and mitigate cognitive and affective symptoms in PD. Finally, our framework is adaptable to other basal ganglia targets and other sleep micro-events (e.g. slow oscillations), providing a versatile platform for sleep neuromodulation in movement disorders and beyond.

Several limitations should be acknowledged. First, while our cohort (n=17) is among the largest synchronized basal ganglia LFP-PSG datasets focused on sleep spindles in PD, the sample size remains modest, particularly for the GPi subgroup (n=4), which precludes robust subregional analysis of this nucleus. Second, all recordings were obtained in the immediate postoperative period with externalized leads; chronic, implanted sensing may exhibit different signal characteristics due to glial encapsulation and long-term stimulation effects. Third, while meta-learning substantially improved cross-subject generalizability, residual inter-individual variability remains, and performance may be influenced by disease severity, dopaminergic medication status, and sleep fragmentation. Fourth, our connectomic analyses relied on a normative connectome rather than patient-specific data, which may limit precision in individuals with advanced neurodegeneration. Future work should validate our pipeline in larger, prospective cohorts, incorporate chronic implanted sensing devices, and evaluate the clinical efficacy of real-time closed-loop spindle modulation in clinical trials.

In conclusion, our study establishes a high-performing, cross-subject compatible pipeline for both the detection and anticipatory prediction of sleep spindles using single-channel basal ganglia LFP signals, representing the first demonstration of such decoding exclusively from subcortical signals. These advances lay the essential preclinical groundwork for the development of spindle-targeted adaptive DBS therapies, with the ultimate goal of restoring physiological sleep architecture and reducing the non-motor symptom burden in Parkinson’s disease.

## Methods

### Study Participants

A total of seventeen patients with idiopathic PD (8 males; mean age 57.2 ± 12.0 years) who had undergone surgical implantation of DBS electrodes targeting the subthalamic nucleus (STN; n = 13) or globus pallidus internus (GPi; n = 4) were enrolled in this retrospective study at Beijing Tiantan Hospital. Participants met the UK Parkinson’s Disease Society Brain Bank diagnostic criteria and exhibited Hoehn and Yahr stages 2–3 in the off-medication state.

Exclusion criteria comprised inability to comply with overnight polysomnography, presence of structural cerebral lesions on preoperative magnetic resonance imaging (MRI), and other major neurological or psychiatric comorbidities. The study protocol was approved by the Institutional Review Board of Beijing Tiantan Hospital (HX-A-2021006) and conducted in accordance with the Declaration of Helsinki. Written informed consent was obtained from all participants prior to study procedures. Detailed demographic and clinical characteristics, including disease duration and levodopa equivalent daily dose, are summarized in Table 1.

### Surgical Procedure and Electrode Localization

Bilateral DBS leads (Medtronic Model 3389 for STN targeting; Model 3387 for GPi targeting) were implanted using standard frame-based stereotactic surgery under local anesthesia, with intraoperative microelectrode recording guidance. Target coordinates were defined by direct visualization on preoperative MRI fused with the Schaltenbrand-Wahren atlas.

Postoperative lead localization was performed using Lead-DBS software (version 3.1) (39). High-resolution postoperative computed tomography (CT) scans (0.625 mm slice thickness) were co-registered with preoperative T1-weighted MRI using advanced normalization tools, followed by nonlinear normalization to the *MNI_ICBM_2009b_NLIN_Asym* standard space. This enabled precise reconstruction of individual electrode contact positions, with mean localization error < 1 mm, supporting subsequent anatomical and connectomic analyses.

### MRI Acquisition

Preoperative MRI scans were acquired on a 3T SIGNA scanner (GE Healthcare) equipped with a 16-channel head coil. Patients were positioned supine with foam padding for head stabilization and ear protection to reduce scanner noise. High-resolution T1-weighted structural images were acquired using a 3D magnetization-prepared rapid gradient-echo (MPRAGE) sequence with the following parameters: repetition time = 9.4 ms, echo time = 4.3 ms, slice thickness = 1 mm, no inter-slice gap, field of view = 256 × 256 mm, matrix = 256 × 256. Resting-state functional MRI data were also collected with eyes closed but were reserved for separate analyses. Postoperative CT scans were acquired within 48 hours after surgery to ensure accurate reconstruction of lead positions.

### Electrophysiological Recordings and Sleep Staging

Sleep recordings were performed in a dedicated sleep laboratory over 1–2 consecutive nights during the lead externalization period, 2–5 days after DBS electrode implantation. Throughout the entire recording session, DBS stimulation was maintained in the off state to eliminate stimulation-related artifacts, and a standardized medication withdrawal protocol was implemented to avoid pharmacologic confounding of neural electrophysiological signals: anti-dystonia medications were discontinued after midday on the recording day, and all anti-parkinsonian medications were stopped after 18:00. Simultaneous whole-night PSG and bilateral basal ganglia LFP signals were acquired via a JE-212 amplifier (Nihon Kohden, Tokyo, Japan). All signals were pre-amplified ×195, hardware bandpass filtered between 0.08 and 300 Hz, and digitized at a sampling rate of 1000 Hz or 2000 Hz. The PSG montage included scalp electroencephalography (EEG) from frontal (F3/F4), central (C3/C4), and occipital (O1/O2) derivations in accordance with the international 10–20 system, as well as bilateral electrooculography, submental electromyography, and synchronized video recording to support standardized sleep staging and artifact rejection. All signal acquisition procedures followed a standardized, clinically validated protocol consistent with the previous workflows (8, 40).

For LFP recordings, bipolar montages were generated from adjacent contacts of each bilateral DBS lead (contact pairs 0–1, 1–2, and 2–3; Figure 1b) targeting either the STN or GPi, with acquisition parameters fully matched to the concurrent PSG signals: standardized ×195 amplification gain, 0.08–300 Hz hardware bandpass filtering, and digitization at a sampling rate of 1000 Hz or 2000 Hz. Sleep stages were manually scored in 30-second epochs by a certified sleep technician blinded to LFP data, following the 2020 American Academy of Sleep Medicine (AASM) criteria. Only non-rapid eye movement (NREM) sleep stages N2 and N3 were retained for subsequent sleep spindle analyses, providing standardized macroarchitectural ground truth for all downstream processing.

### Data Preprocessing

Raw LFP and EEG signals were processed offline with EEGLAB (version 2024.2). A zero-phase fourth-order Butterworth bandpass filter (1–50 Hz) was applied to attenuate low-frequency drifts and high-frequency noise, complemented by a notch filter at 50 Hz (Q = 30) to suppress line noise artifacts. Signals were downsampled to 200 Hz to reduce computational load while preserving oscillatory content of interest. Artifacts arising from patient motion, electrode drift, or amplifier saturation were identified through visual inspection and automated thresholding: epochs exceeding 5 standard deviations of the mean amplitude envelope (computed via Hilbert transform) were excluded. This pipeline yielded artifact-free NREM epochs totaling 4–6 hours per participant, ensuring high-fidelity data for downstream analyses.

### Sleep Spindle Detection and LFP Segment Extraction

Sleep spindles were automatically detected on artifact-free central (C3/C4) and frontal (F3/F4) EEG channels using the YASA toolbox (version 0.6.3) (41). EEG signals were bandpass-filtered (11–16 Hz) using a finite impulse response filter, and root-mean-square (RMS) envelopes were computed with a 0.5-second Hanning window. A spindle event was defined as a segment with RMS amplitude exceeding 1.5 times the mean RMS of all N2/N3 epochs, duration of 0.5–2.0 seconds, and concurrent presence on at least two EEG channels. The temporal midpoint of each validated spindle was used as the reference time stamp.

Corresponding 2-second LFP segments centered on the spindle midpoint were extracted and labeled as *spindle-positive*. To maintain class balance, an identical number of *non-spindle* LFP segments were randomly sampled from 30-second NREM epochs without detectable spindles, divided into 15 non-overlapping 2-second windows, yielding a balanced dataset of approximately 500–1000 samples per class per subject.

### Connectomics-guided Optimal LFP Channel Selection

To automatically identify the optimal LFP channel for spindle decoding in unseen patients, we developed a functional connectomics-guided channel selection framework implemented within a strict leave-one-subject-out (LOSO) cross-validation scheme. Functional connectomics derived from resting-state functional MRI (rs-fMRI) was used in preference to structural tractography due to its higher sensitivity to subtle topographic shifts in local network organization (19).

The framework consisted of three steps:

**1. Functional fingerprint generation**: For each bipolar LFP channel, the volume of tissue activated (VTA) was modeled as a seed region. Voxel-wise Pearson correlations between the VTA seed and whole-brain resting-state BOLD signals were computed using a normative functional connectome from 74 PD patients in the Parkinson’s Progression Markers Initiative (PPMI) database (19, 42), generating a whole-brain functional spatial map (fingerprint) for each channel (see Supplementary Figure 1a).
**2. Group-level representation map (R-map) construction**: Within each training cohort (N-1 subjects), the decoding performance of each channel was evaluated using a robust feature engineering pipeline extracting 10 electrophysiological features: relative spectral power in theta, sigma, high beta, and low gamma bands; low-to-high power ratio (LHPR); sigma-to-beta ratio (SBR); standard deviation of the narrow-band (10–16 Hz) Hilbert envelope; and three Hjorth parameters (activity, mobility, complexity). Features were standardized and fed into an L2-penalized logistic regression classifier with balanced class weights (21). Channels with the highest and lowest balanced accuracies were defined as optimal and worst channels, respectively. Individual difference maps were computed by subtracting the fingerprint of the worst channel from that of the optimal channel (43). A voxel-wise one-sample t-test across all training subjects produced a consensus group-level R-map that isolates the network topology associated with high decoding efficacy (see Supplementary Figure 1b).
**3. Optimal channel assignment**: For each left-out test subject, the functional fingerprint of each available LFP channel was spatially correlated with the training R-map. The channel with the highest R-map match was automatically designated as the optimal channel and used exclusively for all downstream MetaCNN decoding (see Supplementary Figure 1c). The channel with the lowest R-map match was retained for comparative validation.

### Meta-Learning Framework for Cross-Subject Spindle Decoding

To decode spindle events directly from single-channel LFP signals selected by the connectomics pipeline, we developed a meta-learning convolutional neural network (MetaCNN) based on the model-agnostic meta-learning (MAML) paradigm (44). Unlike conventional models limited by inter-subject variability, MetaCNN optimizes a generalizable parameter initialization, enabling fast and robust adaptation to unseen patients using only a minimal calibration dataset (few-shot support set).

Input 2-second LFP segments (400 samples at 200 Hz) were transformed into 2D tensors: a 250-point sliding window with 22-sample steps produced 7 overlapping sub-segments stacked as channels, forming inputs of dimension 7 × 250 × 1. The architecture included three 2D convolutional blocks (3 × 3 kernel, stride 1, batch normalization, exponential linear unit activation), a global average pooling layer, and a fully connected softmax layer for binary classification (spindle versus non-spindle).

During the *meta-representation learning phase*, the model was trained on a series of ‘meta-tasks’ (episodes). Each episode was constructed by sampling LFP data from a small, random group of subjects from the training set, thereby creating diverse and challenging decoding scenarios. Within each episode *K*, the model first performed a temporary, inner-loop update by training its global classifier *θ* on a ‘support set’ (Δ*_kb_*) using cross-entropy loss function Λ*_kb_*.

This produced a set of temporary, task-specific adapted parameters 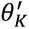 via gradient descent as follows.

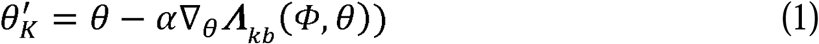

where *Φ* is the parameters of the feature extractor, and α is the inner-loop learning rate.

Crucially, 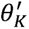 serves merely as a temporary adapted state simulating the personalization process for the specific decoding scenario *K*. The overarching algorithmic objective is not to retain 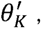 but to utilize its performance feedback to optimize the adaptable global initialization (*Φ* and *θ*). To this end, in the subsequent outer loop, the model evaluated these temporarily adapted parameters on a separated ‘query set’ (Δ*_km_*) to compute the meta-loss *Λ_km_*. The gradient of the meta-loss was then backpropagated all the way to the original global parameters, performing a meta-update driven by an Adam optimizer.

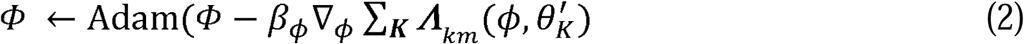

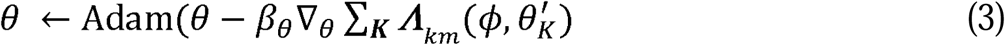

where *β_Φ_* and *β_θ_* represent the differential outer-loop learning rates, guided by a step-wise learning rate scheduler (see Table 2 for details).

During the *meta-adaptation phase* (i.e., the testing step of the LOSO procedure), the meta-trained model was evaluated on the new, held-out subject. As depicted in Figure 2a and Supplementary Figure 3, the model used a small ‘support set’ of labeled examples from the new subject to generate a subject-specific “task guide.” This guide effectively creates a temporary, adapted classifier state, which was then used to decode the remaining spindle and non-spindle segments (the query set) from that subject. Training proceeded in episodes sampling tasks from the distribution of subjects, with LOSO cross-validation (train on N-1 subjects, test on held-out), as well. To validate clinical feasibility for closed-loop neuromodulation, we measured two key latency metrics: (1) meta-adapt latency for online calibration using the few-shot support set; (2) real-time inference latency for processing a single 2-second query frame.

### Benchmark Decoding Models

MetaCNN performance was benchmarked against two standard approaches under identical LOSO protocols. First, a feature-based support vector machine (SVM; *scikit-learn* package version 1.7.2, radial basis function kernel) was trained based on two physically interpretable features extracted from the selected single-channel LFP: peak power and phase-locking value (PLV). Specifically, to strictly quantify local activity without the confounding effect of aperiodic background noise (1/f), we applied the ‘specparam’ (FOOOF) algorithm to parameterize the neural power spectra of LFP segments, isolating the pure peak power within the sigma band (11–16 Hz) (45). Furthermore, to evaluate the network-level phase synchronization between the basal ganglia and the cortex, we computed the PLV between the selected deep LFP channel and the corresponding scalp EEG channel (C3/C4). The PLV was calculated precisely in the time-frequency domain using the continuous wavelet transform (CWT) with complex Morlet wavelets (45). Hyperparameters C and γ were tuned via grid search per fold.

Second, we employed a base convolutional neural network (BaseCNN) (46, 47) that learns features directly from the raw LFP signal in an end-to-end fashion. The BaseCNN model is composed of a feature extractor followed by a final classification layer. The feature extractor consists of three convolutional blocks, each containing a 2D convolutional layer, a batch normalization layer, and an Exponential Linear Unit (ELU) activation function, designed to capture non-linearly hierarchical features from the LFP time-series data. The final classification layer is a fully-connected linear layer with a softmax output for the binary classification of spindle versus non-spindle segments. The model was trained using a standard Adam optimizer. The detailed architecture and training hyperparameters are provided in Supplementary Table 3.

All models were evaluated using four standard metrics: accuracy, F1-score, sensitivity, and specificity.

### Spatial and Electrophysiological Mechanism Analysis

To characterize the neural and anatomical substrates underlying connectomics-guided channel selection, we performed multi-modal validation:

**1. Selection strategy validation**: MetaCNN decoding accuracy was compared between channels with the highest R-map match and those with the lowest R-map match within the same subjects.
**2. Electrophysiological characterization**: FOOOF-derived sigma-band peak power and CWT-derived PLV were compared between spindle and non-spindle LFP segments to quantify local and network-level spindle-related modulation.
**3. Anatomical mapping**: Electrode contacts were localized and projected onto 3D surface models of the STN and GPi in MNI space. Decoding performance was z-scored and spatially mapped to identify efficacy hotspots. For STN subregional parcellation, the DISTAL atlas was used to define motor, associative, and limbic subdivisions. Spherical VTAs (radii 1.0–10.0 mm in 0.5 mm increments) were modeled, and intra-subject overlap volume differences (ΔVolume) between highest and lowest R-map matched channels were quantified for each STN subregion. Overlap with the limbic STN subregion at a standardized 4-mm radius was statistically tested.

### Predictive Spindle Decoding and Temporal Horizon Analysis

To evaluate anticipatory prediction of sleep spindles from basal ganglia LFPs, we performed a temporal horizon analysis without modifying the MetaCNN architecture. A 2-second LFP observation window was systematically shifted backward relative to cortical spindle onset, with prediction horizon (Δt) defined as the interval between the end of the predictive window and spindle onset (Δt = 0.0 s to −2.0 s). Segments immediately preceding spindle onset were labeled *pre-spindle-positive*. Class-balanced non-spindle segments were sampled equivalently from spindle-free NREM epochs. The identical MetaCNN, SVM, and BaseCNN architectures were re-applied under LOSO cross-validation to assess cross-subject anticipatory decoding performance across progressive temporal horizons.

### Statistical Analysis

All statistical analyses were performed in Python (version 3.10) using SciPy and Pingouin libraries. Normality was assessed using the Shapiro-Wilk test. Paired group comparisons were performed using two-tailed paired t-tests (normally distributed data) or Wilcoxon signed-rank tests (non-parametric data). For temporal horizon analyses involving multiple comparisons, P-values were adjusted using the Bonferroni correction to control family-wise error rate. Associations between VTA overlap volumes and decoding accuracy were assessed using Spearman rank correlations. Statistical significance was defined as (\**p* < 0.05, \*\**p* < 0.01, \*\*\**p* < 0.001). Data are presented as mean ± standard deviation (SD) or median with interquartile range (IQR) as appropriate.

## Acknowledgments

This study is supported by grants from National Natural Science Foundation of P.R. China (62106113, 62276081, 82571664), Guangdong Basic and Applied Basic Research Foundation (2023A1515010792, 2023B1515120065), Shenzhen Science and Technology Program (GXWD20231129121139001, JCYJ20240813110522029), the Beijing Natural Science Foundation (7264270).

## Data availability

The data collected in this study are available from the corresponding author, upon reasonable request.

## Code availability

Statistical analyses were performed in R. Code for all analyses and visualizations is released in https://github.com/chenfei-ye/spindle_LFP_decoding.

## Author Contributions

C.Y. and Z.Y. conceptualized this work. J.L. and C.Y. developed the analytical methods, performed data analysis, prepared the figures, and drafted the manuscript. Z.Y. led clinical data collection and co-performed data analysis. J.L. completed the preprocessing of electrophysiological data. Y.L., Y.X., and H.F. participated in clinical data collection and electrophysiological data preprocessing. T.M. and J.Z. provided overall supervision for this study.

