## Supplementary File for "Connectomics-guided meta-learning for decoding and anticipatory prediction of sleep spindles from basal ganglia local field potentials in Parkinson’s disease"

**Single-Channel Basal Ganglia LFP Enables Cross-Subject Sleep Spindle Decoding**

**Supplementary File**

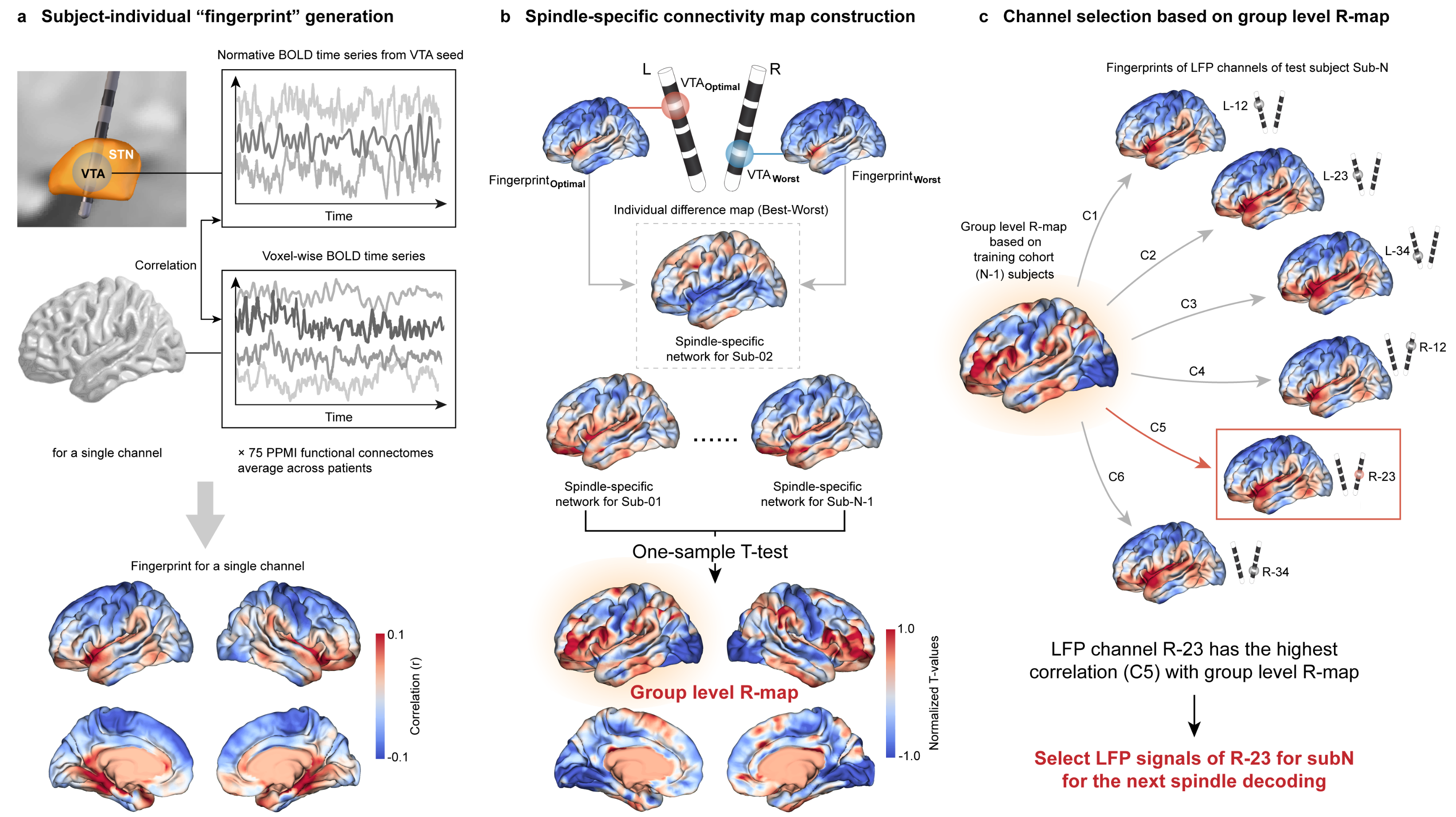

**Supplementary Figure 1 Connectomics-guided framework identifies individualized LFP channels for cross-subject spindle decoding. a.** Generation of subject-specific functional connectivity "fingerprints". Normative resting-state functional MRI data ((n = 74 PPMI PD patients) were used to compute the correlation between the volume of tissue activated (VTA) seed of each bipolar LFP channel and whole-brain voxels. **b**. Construction of the spindle-specific group-level representation map (R-map). Individual difference maps were calculated by contrasting the fingerprints of the highest and lowest performing channels. A one-sample t-test across the training cohort (N-1 subjects) yielded the consensus Group-level R-map. **c.** Leave-one-subject-out cross-validation strategy for optimal channel selection. For a new test subject, the functional fingerprint of each available LFP channel is spatially correlated with the Group-level R-map. The channel exhibiting the highest spatial correlation (e.g., R-23) is automatically selected for downstream spindle decoding.

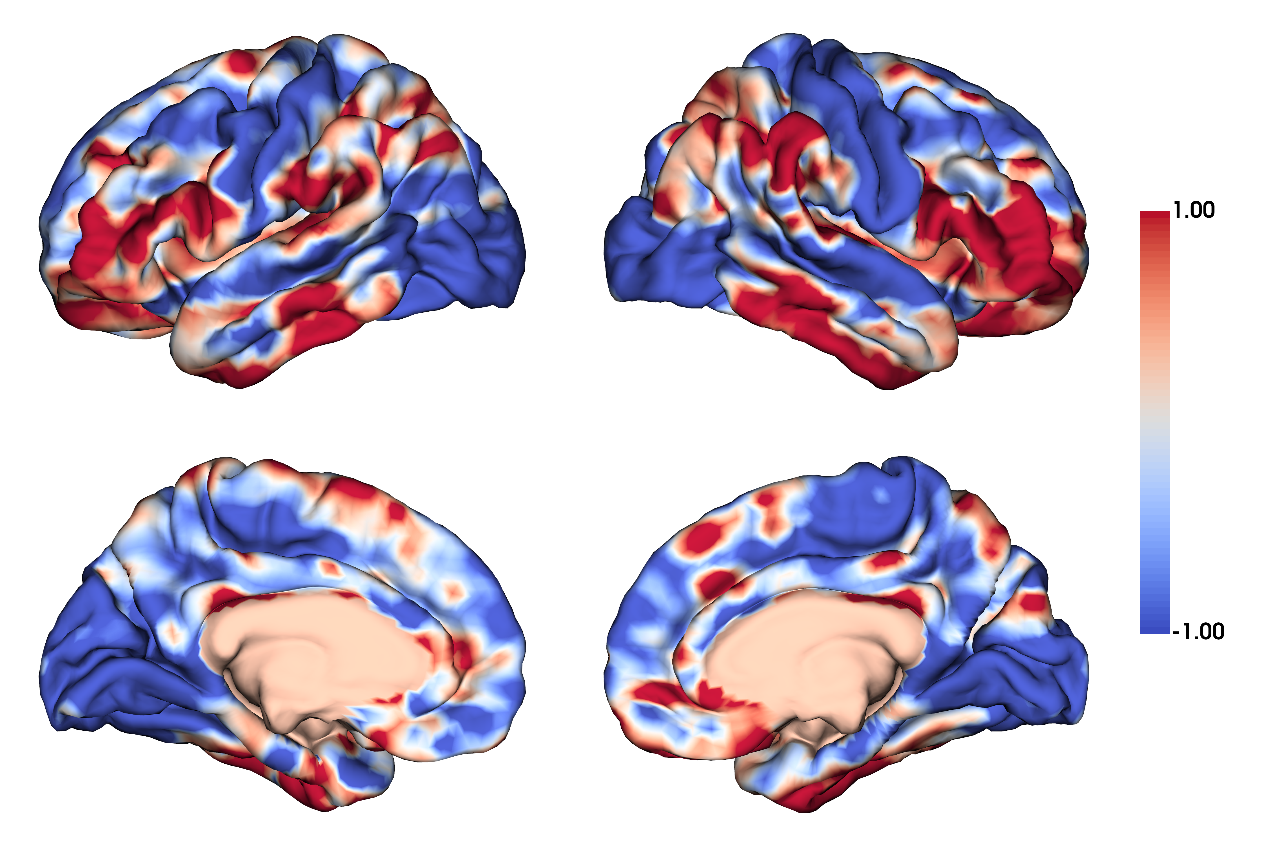

**Supplementary Figure 2 Consensus group-level representation map (R-map) of the brain-wide anatomical network driving spindle decoding efficacy.** The topography was generated utilizing the functional connectivity fingerprints from the entire patient cohort, delineating a highly polarized network topology. High spindle decoding performance is positively associated (hot colors) with connectivity to prefrontal and limbic hubs, primarily localized within the orbitofrontal and anterior cingulate cortex. Conversely, strong negative connectivity (cold colors) is linked to suboptimal decoding efficacy, notably within primary sensorimotor areas and the superior parietal cortex. The color bar represents the normalized correlation values ranging from -1.00 to 1.00 (peak raw T-statistics: T_max_ = 4.46, T_min_ = -4.08, both *P* < 0.001).

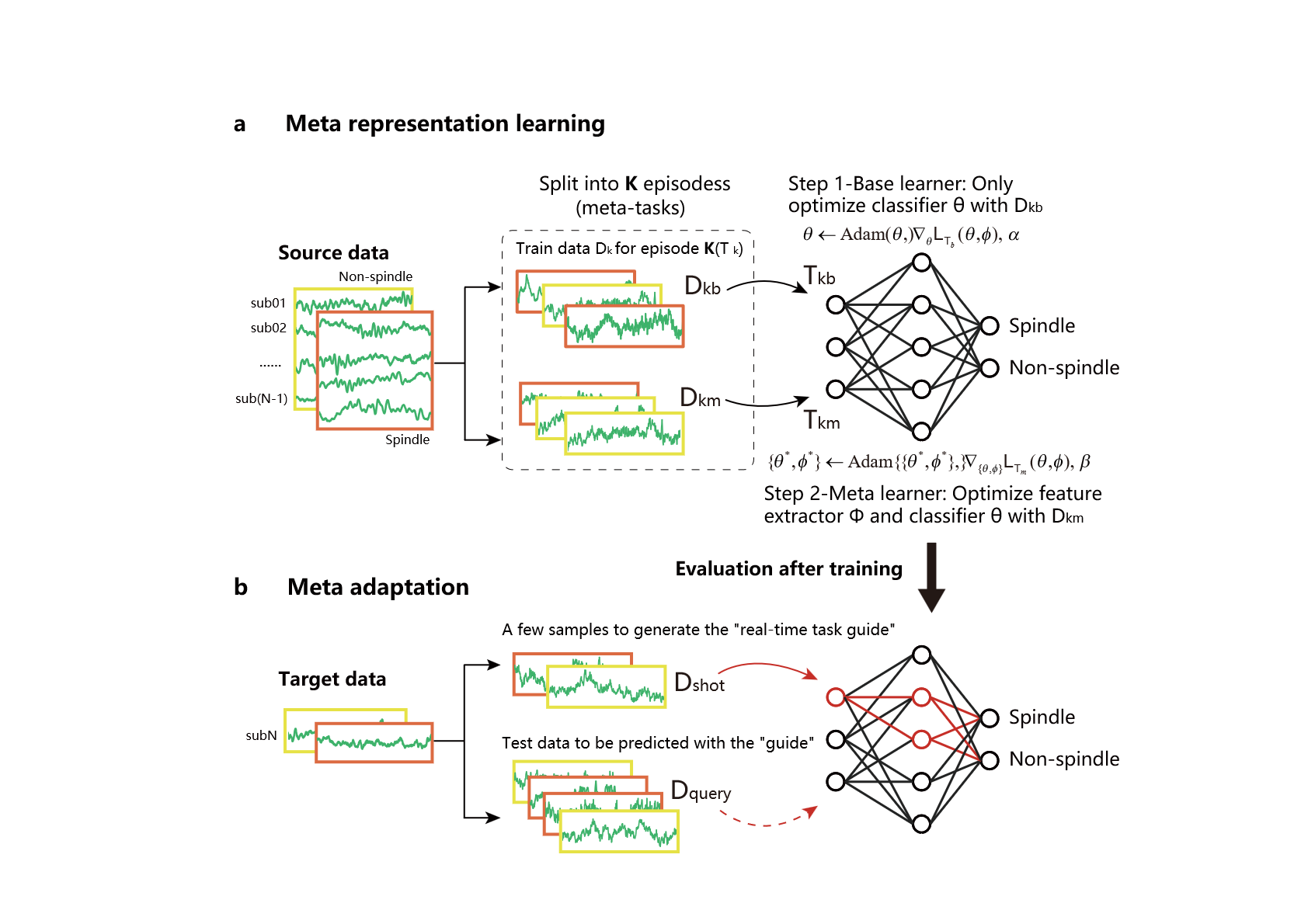

**Supplementary Figure 3** **Meta-learning framework for cross-subject spindle decoding**. **a** The meta-representation learning phase trains the model to find an optimal initialization for rapid adaptation. The model learns across numerous 'meta-tasks' (episodes), each constructed by sampling LFP data from a small group of source subjects to create diverse, challenging decoding scenarios. Within each episode, the model uses a 'support set’ (D_kb_) of examples to perform a temporary update, simulating adaptation to this specific data mix. It then refines its core feature extractor and classifier based on its performance on a separate 'query set’ (D_km_). This process optimizes for a generalized feature representation that is robust to inter-subject variability. **b** In the meta-adaptation phase, the trained model is evaluated on a new, unseen target subject. Using only a few labeled examples from the new subject (support set, D_shot_), the model generates a subject-specific "task guide" that modulates its feature processing in a single forward pass. This allows for accurate spindle decoding on the test data (query set, D_query_) without requiring any further weight updates or retraining. Red connections illustrate this dynamic, context-specific pathway.

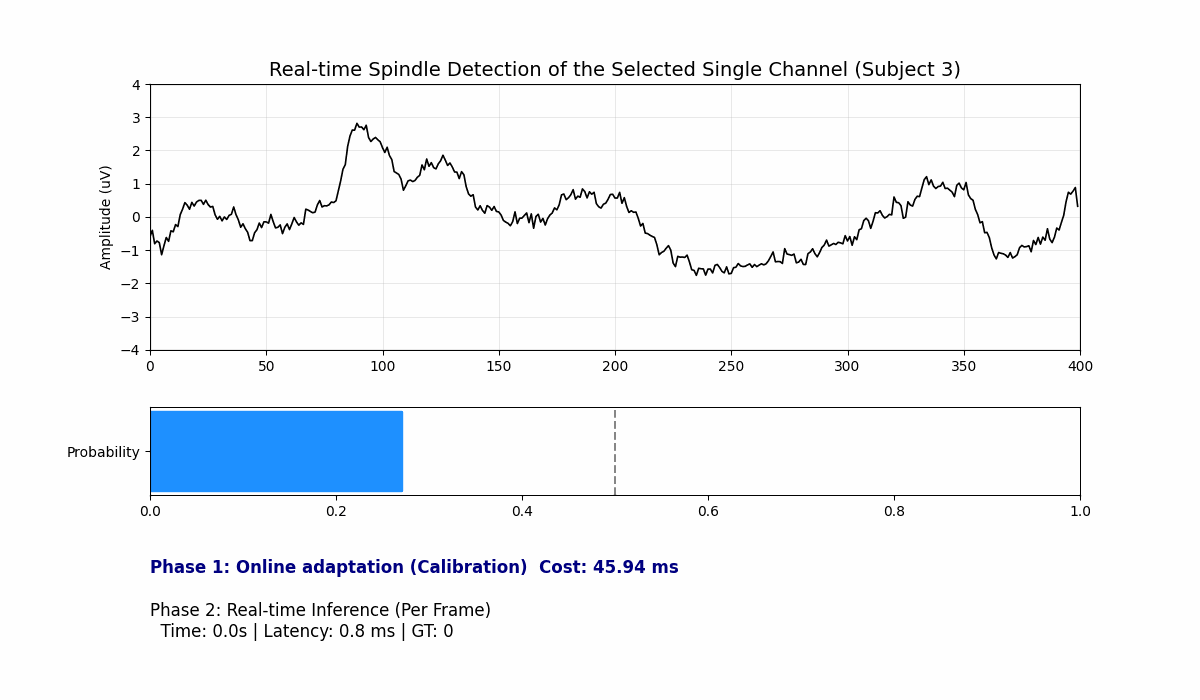

**Supplementary Figure 4** **Snapshot of dynamic real-time spindle decoding during continuous LFP streaming.** This visualization corresponds to the real-time inference phase (query set data stream) illustrated in Figure 3a, simulating closed-loop detection utilizing the optimal selected single channel from a representative test patient (Subject 3). **Top panel:** A 2-second sliding window of the continuous LFP signal. The dynamic background explicitly highlights the current detection status, with green indicating a True Positive (TP) spindle detection. **Middle panel:** The real-time output probability generated by the MetaCNN classifier. The vertical dashed line represents the decision threshold (0.5). **Bottom panel:** Real-time computational tracking metrics. The initial patient-specific meta-adaptation (calibration) required 45.94 ms, while the continuous real-time inference achieved a latency of ~0.9 ms per query frame.

| **Supplementary Table 1** Individual participant statistics of detected sleep spindle events during NREM sleep | | | | | | | | |
| --- | --- | --- | --- | --- | --- | --- | --- | --- |
| **Patients** | **Num. of spindles** | **Num. of spindles in N2** | **Num. of spindles in N3** | **Ratio of spindles in N2**  **(%)** | **Ratio of spindles in N3**  **(%)** | **Avg. RMS fold** | **Ratio of Frontal spindles (%)** | **Avg. Freq. of Frontal spindles**  **(Hz)** |
| *STN-Lead* | | | | | | | | |
| STN004 | 277 | 277 | 0 | 100.00 | 0.00 | 2.62 | 71.96 | 13.01 |
| STN010 | 155 | 151 | 4 | 97.42 | 2.58 | 2.65 | 85.43 | 12.88 |
| STN011 | 159 | 158 | 1 | 99.37 | 0.63 | 2.61 | 76.10 | 12.66 |
| STN063 | 131 | 131 | 0 | 100.00 | 0.00 | 2.76 | 81.75 | 12.25 |
| STN070 | 151 | 149 | 2 | 98.68 | 1.32 | 2.78 | 84.56 | 12.57 |
| STN084 | 204 | 204 | 0 | 100.00 | 0.00 | 2.42 | 20.87 | 13.15 |
| STN090 | 292 | 273 | 19 | 93.49 | 6.51 | 2.13 | 85.27 | 12.85 |
| STN112 | 283 | 283 | 0 | 100.00 | 0.00 | 2.79 | 37.53 | 13.46 |
| STN116 | 190 | 188 | 2 | 98.95 | 1.05 | 2.58 | 72.04 | 12.13 |
| STN117 | 158 | 142 | 16 | 89.87 | 10.13 | 2.74 | 37.42 | 13.12 |
| STN143 | 151 | 149 | 2 | 98.68 | 1.32 | 2.40 | 78.67 | 13.19 |
| STN150 | 274 | 273 | 1 | 99.64 | 0.36 | 2.55 | 81.85 | 12.96 |
| STN171 | 193 | 193 | 0 | 100.00 | 0.00 | 2.39 | 74.49 | 12.52 |
| *GPi-Lead* | | | | | | | | |
| GPi001 | 140 | 137 | 3 | 97.86 | 2.14 | 2.66 | 92.91 | 13.16 |
| GPi071 | 424 | 410 | 14 | 96.70 | 3.30 | 2.26 | 96.50 | 12.49 |
| GPi108 | 146 | 137 | 9 | 93.84 | 6.60 | 3.06 | 93.12 | 13.27 |
| GPi153 | 108 | 98 | 10 | 90.74 | 9.26 | 3.04 | 52.78 | 13.01 |
| **Avg.** | 202.12 | 197.24 | 4.88 | 97.37 | 2.66 | 2.61 | 71.96 | 12.86 |

*Num.* Number. *Avg.* Average. *RMS* Root mean square. *Freq.* Frequency.

| **Supplementary Table 2** Subject-wise spatial correlations of LFP functional fingerprints to the group-level R-map | | | | |
| --- | --- | --- | --- | --- |
| **Patients** | **Correlation of highest R-map match** | **Channel with highest R-map match (Selected)** | **Correlation of lowest R-map match** | **Channel with lowest R-map match** |
| *STN-Lead* | | | | |
| STN004 | 0.253 | L-12 | 0.217 | L-34 |
| STN010 | 0.137 | L-12 | 0.068 | R-34 |
| STN011 | 0.184 | R-34 | 0.127 | L-23 |
| STN063 | 0.149 | R-34 | 0.081 | L-23 |
| STN070 | 0.150 | R-12 | 0.051 | L-23 |
| STN084 | 0.142 | L-12 | 0.095 | R-23 |
| STN090 | 0.175 | L-12 | 0.120 | R-23 |
| STN112 | 0.167 | L-34 | 0.092 | R-34 |
| STN116 | 0.131 | L-12 | 0.035 | R-23 |
| STN117 | 0.182 | L-12 | 0.119 | L-34 |
| STN143 | 0.108 | L-12 | 0.028 | L-34 |
| STN150 | 0.119 | L-12 | 0.065 | L-34 |
| STN171 | 0.097 | R-12 | 0.005 | L-23 |
| *GPi-Lead* | | | | |
| GPi001 | 0.104 | L-34 | -0.110 | L-12 |
| GPi071 | 0.186 | L-34 | 0.023 | L-12 |
| GPi108 | 0.268 | L-34 | -0.054 | R-12 |
| GPi153 | 0.093 | R-34 | -0.060 | R-12 |
| **Avg.** | 0.156 | - | 0.053 | - |

*R-map* Representation map*. L-12* Left channel-12, *L-23* Left channel-23, *L-34* Left channel-34, *R-12* Right channel-12, *R-23* Right channel-23, *R-34* Right channel-34.

| **Supplementary Table 3** Detailed architecture and training parameters of the CNN models | | | | |
| --- | --- | --- | --- | --- |
| **Part A**. Network architecture of the CNN models | | | | |
| Component | Layer | Type | Details | Output shape |
| Input data |  | 1D LFP segment | Single channel, 2 seconds @ 200 Hz | (1, 400) |
| Data transformation |  | Sliding window | Window: 250 time points,  Step: 22 time points | 7 windows of (250, 1) |
| Model Input |  | 2D feature map | 7 stacked temporal windows | (7, 250, 1) |
| Feature extractor | Block 1 | Conv2D + BatchNorm2D | Kernel: (22, 1), Stride: 1,  Filters: 16 | (16, 229, 1) |
|  |  | Activation | ELU |  |
|  | Block 2 | Conv2D + BatchNorm2D | Kernel: (2, 32), Stride: 1,  Filters: 4 |  |
|  |  | Activation | ELU |  |
|  |  | MaxPool2D | Kernel: (2, 4), Stride: 4 | (4, H''/2, W''/4) |
|  | Block 3 | Conv2D + BatchNorm2D | Kernel: (8, 4), Stride: 1,  Filters: 4 | (4, H''', W''') |
|  |  | Activation | ELU |  |
|  |  | MaxPool2D | Kernel: (2, 4), Stride: 4 | (4, H''''/2, W''''/4) |
|  | Flatten | Reshape | - | 112 |
| Classifier | Output layer | Fully connected layer | Neurons: 2 | 2 |
|  |  | Activation | Softmax |  |

ELU denotes the Exponential Linear Unit activation function. Data transformation, using a sliding window, converts the 1D LFP time-series into a 2D feature map with 7 channels to serve as the model input. Padding was applied before the second and third convolutional layers.

| **Part B**. Training parameters of the CNN models | | | |
| --- | --- | --- | --- |
| Parameter | | BaseCNN | MetaCNN |
| Optimizer | | Adam | Adam |
| Learning rate | Feature extractor | 1e-4 | 1e-4 |
|  | Classifier | 1e-4 | 5e-3 |
| Learning rate scheduler | | - | Step size: 3,  Decay rate: 0.8 |

| **Supplementary Table 4** Subject-wise single-channel concurrent decoding and anticipatory predictive decoding performance of MetaCNN, using the LFP channel with the highest R-map match. | | | | | | | | | |
| --- | --- | --- | --- | --- | --- | --- | --- | --- | --- |
| **Patients** | **Channel Selected** | **Decoding performance** | | | | **Predictive decoding performance** | | | |
|  |  | **ACC** | **Sen** | **Spe** | **F1** | **ACC** | **Sen** | **Spe** | **F1** |
| *STN-Lead* | | | | | | | | | |
| STN004 | L-12 | 93.25 | 91.50 | 95.00 | 93.04 | 88.25 | 84.50 | 92.00 | 87.59 |
| STN010 | L-12 | 95.50 | 93.00 | 98.00 | 95.28 | 85.00 | 84.00 | 86.00 | 84.76 |
| STN011 | R-34 | 89.25 | 80.50 | 98.00 | 87.74 | 80.25 | 81.00 | 79.50 | 80.42 |
| STN063 | R-34 | 96.75 | 93.50 | 100.00 | 96.43 | 83.25 | 82.50 | 84.00 | 83.35 |
| STN070 | R-12 | 90.25 | 84.50 | 96.00 | 89.34 | 85.50 | 81.50 | 89.50 | 84.44 |
| STN084 | L-12 | 92.75 | 85.50 | 100.00 | 91.75 | 82.00 | 80.50 | 83.50 | 81.42 |
| STN090 | L-12 | 95.75 | 96.00 | 95.50 | 95.76 | 87.50 | 94.00 | 81.00 | 88.38 |
| STN112 | L-34 | 89.50 | 87.50 | 91.50 | 89.28 | 83.50 | 88.00 | 79.00 | 84.42 |
| STN116 | L-12 | 90.50 | 83.50 | 97.50 | 89.19 | 85.25 | 78.00 | 92.50 | 83.42 |
| STN117 | L-12 | 96.75 | 97.00 | 96.50 | 96.78 | 76.75 | 80.50 | 73.00 | 77.40 |
| STN143 | L-12 | 93.25 | 89.00 | 97.50 | 92.73 | 78.25 | 75.50 | 81.00 | 77.55 |
| STN150 | L-12 | 92.25 | 94.00 | 90.50 | 92.39 | 86.00 | 89.00 | 83.00 | 86.16 |
| STN171 | R-12 | 95.50 | 92.50 | 98.50 | 95.12 | 86.00 | 86.50 | 85.50 | 86.06 |
| *GPi-Lead* | | | | | | | | | |
| GPi001 | L-34 | 96.50 | 93.00 | 100.00 | 96.23 | 83.75 | 85.50 | 82.00 | 83.93 |
| GPi071 | L-34 | 86.50 | 89.00 | 84.00 | 86.59 | 74.75 | 75.50 | 74.00 | 74.66 |
| GPi108 | L-34 | 90.50 | 89.50 | 91.50 | 90.38 | 87.25 | 82.50 | 92.00 | 86.57 |
| GPi153 | R-34 | 90.00 | 87.00 | 93.00 | 89.22 | 86.50 | 94.00 | 79.00 | 87.58 |
| **Avg** | | 92.63 | 89.79 | 95.47 | 92.19 | 83.44 | 83.42 | 83.47 | 83.30 |
| **Std** | | 3.13 | 4.61 | 4.24 | 3.32 | 3.92 | 5.44 | 5.83 | 3.13 |

*L-12* Left channel-12, *L-23* Left channel-23, *L-34* Left channel-34, *R-12* Right channel-12, *R-23* Right channel-23, *R-34* Right channel-34. *Avg* Average. *Std* Standard deviation. *ACC* accuracy, *Sen* sensitivity, *Spe* specificity, *F1* F1-score.

| **Supplementary Table 5** Subject-wise single-channel concurrent decoding and anticipatory predictive decoding performance of BaseCNN, using the LFP channel with the highest R-map match. | | | | | | | | | |
| --- | --- | --- | --- | --- | --- | --- | --- | --- | --- |
| **Patients** | **Channel Selected** | **Decoding performance** | | | | **Predictive decoding performance** | | | |
|  |  | **ACC** | **Sen** | **Spe** | **F1** | **ACC** | **Sen** | **Spe** | **F1** |
| *STN-Lead* | | | | | | | | | |
| STN004 | L-12 | 81.18 | 69.74 | 92.62 | 78.75 | 51.85 | 47.41 | 56.30 | 49.61 |
| STN010 | R-34 | 71.33 | 71.33 | 71.33 | 71.33 | 45.16 | 47.10 | 43.23 | 46.20 |
| STN011 | L-12 | 79.69 | 75.00 | 84.38 | 78.69 | 48.73 | 54.78 | 42.68 | 51.65 |
| STN063 | R-23 | 73.13 | 78.36 | 67.91 | 74.47 | 53.68 | 54.41 | 52.94 | 54.01 |
| STN070 | R-34 | 64.47 | 72.37 | 56.58 | 67.07 | 48.69 | 49.02 | 48.37 | 48.86 |
| STN084 | L-12 | 83.25 | 75.94 | 90.57 | 81.93 | 52.14 | 55.71 | 48.57 | 53.79 |
| STN090 | L-34 | 85.14 | 80.41 | 89.86 | 84.40 | 47.57 | 47.92 | 47.22 | 47.75 |
| STN112 | L-12 | 84.16 | 75.09 | 93.24 | 82.58 | 50.00 | 49.82 | 50.18 | 49.91 |
| STN116 | R-12 | 72.75 | 65.08 | 80.42 | 70.49 | 46.83 | 43.92 | 49.74 | 45.23 |
| STN117 | L-12 | 79.01 | 76.54 | 81.48 | 78.48 | 54.82 | 59.04 | 50.60 | 56.65 |
| STN143 | R-12 | 82.41 | 74.48 | 90.34 | 80.90 | 53.95 | 48.03 | 59.87 | 51.05 |
| STN150 | R-34 | 77.99 | 69.72 | 86.27 | 76.01 | 50.14 | 54.87 | 45.40 | 52.39 |
| STN171 | L-34 | 62.05 | 59.49 | 64.62 | 61.05 | 49.23 | 53.85 | 44.62 | 51.47 |
| *GPi-Lead* | | | | | | | | | |
| GPi001 | R-34 | 69.93 | 79.72 | 60.14 | 72.61 | 48.23 | 45.39 | 51.06 | 46.72 |
| GPi071 | L-34 | 75.60 | 77.18 | 74.03 | 75.98 | 49.88 | 60.00 | 39.77 | 54.49 |
| GPi108 | L-12 | 74.83 | 72.41 | 77.24 | 74.20 | 55.52 | 53.10 | 57.93 | 54.42 |
| GPi153 | L-23 | 78.83 | 71.17 | 86.49 | 77.07 | 54.81 | 48.15 | 61.48 | 51.59 |
| **Avg** | | 76.22 | 73.18 | 79.27 | 75.65 | 50.66 | 51.32 | 50.00 | 50.93 |
| **Std** | | 6.67 | 5.31 | 11.69 | 5.95 | 3.09 | 4.75 | 6.17 | 3.24 |

*L-12* Left channel-12, *L-23* Left channel-23, *L-34* Left channel-34, *R-12* Right channel-12, *R-23* Right channel-23, *R-34* Right channel-34. *Avg* Average. *Std* Standard deviation. *ACC* accuracy, *Sen* sensitivity, *Spe* specificity, *F1* F1-score.

| **Supplementary Table 6** Subject-wise single-channel concurrent decoding and anticipatory predictive decoding performance of the support vector machine (SVM), using the LFP channel with the highest R-map match. | | | | | | | | | |
| --- | --- | --- | --- | --- | --- | --- | --- | --- | --- |
| **Patients** | **Channel Selected** | **Decoding performance** | | | | **Predictive decoding performance** | | | |
|  |  | **ACC** | **Sen** | **Spe** | **F1** | **ACC** | **Sen** | **Spe** | **F1** |
| *STN-Lead* | | | | | | | | | |
| STN004 | L-12 | 72.75 | 92.70 | 52.81 | 77.28 | 53.60 | 72.07 | 35.14 | 60.84 |
| STN010 | L-12 | 78.66 | 73.17 | 84.15 | 77.42 | 55.95 | 83.33 | 28.57 | 65.42 |
| STN011 | R-34 | 73.23 | 90.91 | 55.56 | 77.25 | 50.00 | 69.70 | 30.30 | 58.23 |
| STN063 | R-34 | 72.00 | 60.00 | 84.00 | 68.18 | 57.41 | 88.89 | 25.93 | 67.61 |
| STN070 | R-12 | 67.76 | 39.47 | 96.05 | 55.05 | 51.69 | 76.27 | 27.12 | 61.22 |
| STN084 | L-12 | 68.79 | 95.74 | 41.84 | 75.42 | 53.40 | 57.28 | 49.51 | 55.14 |
| STN090 | L-12 | 55.16 | 99.46 | 10.86 | 68.93 | 57.09 | 61.19 | 52.99 | 58.78 |
| STN112 | L-34 | 78.19 | 81.86 | 74.50 | 78.96 | 41.56 | 63.64 | 19.48 | 52.13 |
| STN116 | L-12 | 81.02 | 84.26 | 77.78 | 81.61 | 57.00 | 76.00 | 38.00 | 63.87 |
| STN117 | L-12 | 65.48 | 39.29 | 91.67 | 53.22 | 42.50 | 80.00 | 5.00 | 58.18 |
| STN143 | L-12 | 80.95 | 78.57 | 83.33 | 80.49 | 47.50 | 75.00 | 20.00 | 58.82 |
| STN150 | L-12 | 62.04 | 29.84 | 94.24 | 44.02 | 49.28 | 74.82 | 23.74 | 59.60 |
| STN171 | R-12 | 64.67 | 40.00 | 89.33 | 53.10 | 53.66 | 82.93 | 24.39 | 64.15 |
| *GPi-Lead* | | | | | | | | | |
| GPi001 | L-34 | 75.00 | 58.57 | 91.43 | 70.09 | 53.13 | 81.25 | 25.00 | 63.41 |
| GPi071 | L-34 | 71.33 | 89.42 | 53.24 | 75.72 | 59.94 | 67.84 | 52.05 | 62.87 |
| GPi108 | L-34 | 73.24 | 57.75 | 88.73 | 68.33 | 48.48 | 72.73 | 24.24 | 58.54 |
| GPi153 | R-34 | 68.49 | 53.42 | 83.56 | 62.90 | 51.47 | 85.29 | 17.65 | 63.74 |
| **Avg** | | 71.10 | 68.50 | 73.71 | 68.70 | 51.98 | 74.60 | 29.36 | 60.74 |
| **Std** | | 6.91 | 22.77 | 23.18 | 11.30 | 5.07 | 8.74 | 12.80 | 3.90 |

*L-12* Left channel-12, *L-23* Left channel-23, *L-34* Left channel-34, *R-12* Right channel-12, *R-23* Right channel-23, *R-34* Right channel-34. *Avg* Average. *Std* Standard deviation. *ACC* accuracy, *Sen* sensitivity, *Spe* specificity, *F1* F1-score.

| **Supplementary Table 7** Subject-wise online adaptation time and inference time of MetaCNN | | | | |
| --- | --- | --- | --- | --- |
| **Patient** | **Adaptation time (ms)** | **Std. of adaptation time (ms)** | **Inference time**  **(****ms/per frame)** | **Std. of**  **inference time**  **(ms/per frame)** |
| STN004 | 45.36 | 1.78 | 0.91 | 0.33 |
| STN010 | 48.20 | 3.81 | 1.06 | 0.39 |
| STN011 | 37.20 | 0.08 | 0.96 | 0.62 |
| STN063 | 38.15 | 5.93 | 0.99 | 0.88 |
| STN070 | 49.48 | 0.25 | 1.06 | 0.39 |
| STN084 | 45.65 | 0.07 | 0.93 | 0.33 |
| STN090 | 45.06 | 4.43 | 0.95 | 0.33 |
| STN112 | 39.06 | 7.55 | 0.95 | 0.34 |
| STN116 | 55.10 | 1.53 | 0.96 | 0.35 |
| STN117 | 49.84 | 1.58 | 1.06 | 0.39 |
| STN143 | 52.69 | 2.92 | 0.97 | 0.34 |
| STN150 | 45.72 | 1.60 | 0.93 | 0.33 |
| STN171 | 44.21 | 1.81 | 0.99 | 0.80 |
| GPi001 | 44.38 | 0.46 | 0.95 | 0.34 |
| GPi071 | 43.49 | 2.64 | 0.94 | 0.33 |
| GPi108 | 41.42 | 3.98 | 0.94 | 0.33 |
| GPi153 | 54.63 | 1.55 | 0.95 | 0.34 |
| **Avg** | 45.86 | 2.47 | 0.97 | 0.42 |

*Avg* Average. *Std* Standard deviation.

| **Supplementary Table 8** Subject-wise single-channel concurrent decoding and anticipatory predictive decoding performance of MetaCNN, using the LFP channel with the lowest R-map match. | | | | | | | | | |
| --- | --- | --- | --- | --- | --- | --- | --- | --- | --- |
| **Patients** | **Channel Selected** | **Decoding performance** | | | | **Predictive decoding performance** | | | |
|  |  | **ACC** | **Sen** | **Spe** | **F1** | **ACC** | **Sen** | **Spe** | **F1** |
| *STN-Lead* | | | | | | | | | |
| STN004 | L-34 | 90.25 | 86.50 | 94.00 | 89.60 | 82.25 | 80.00 | 84.50 | 81.38 |
| STN010 | R-34 | 90.50 | 92.50 | 88.50 | 90.71 | 74.75 | 75.00 | 74.50 | 74.44 |
| STN011 | L-23 | 90.25 | 90.50 | 90.00 | 90.17 | 86.00 | 88.00 | 84.00 | 86.04 |
| STN063 | L-23 | 92.00 | 94.00 | 90.00 | 92.24 | 81.25 | 83.50 | 79.00 | 81.21 |
| STN070 | L-23 | 87.75 | 83.50 | 92.00 | 86.82 | 82.00 | 79.50 | 84.50 | 81.31 |
| STN084 | R-23 | 89.25 | 91.00 | 87.50 | 89.43 | 77.75 | 78.50 | 77.00 | 77.91 |
| STN090 | R-23 | 85.75 | 83.00 | 88.50 | 85.22 | 78.75 | 76.00 | 81.50 | 78.15 |
| STN112 | R-34 | 87.25 | 94.50 | 80.00 | 88.17 | 77.50 | 75.00 | 80.00 | 76.04 |
| STN116 | R-23 | 93.00 | 95.00 | 91.00 | 93.17 | 82.50 | 85.00 | 80.00 | 83.03 |
| STN117 | L-34 | 92.50 | 92.00 | 93.00 | 92.43 | 76.00 | 76.50 | 75.50 | 76.01 |
| STN143 | L-34 | 88.75 | 88.00 | 89.50 | 88.66 | 76.00 | 75.00 | 77.00 | 75.02 |
| STN150 | L-34 | 89.25 | 86.50 | 92.00 | 88.63 | 83.50 | 81.00 | 86.00 | 82.97 |
| STN171 | L-23 | 93.25 | 93.00 | 93.50 | 93.20 | 77.75 | 76.50 | 79.00 | 77.21 |
| *GPi-Lead* | | | | | | | | | |
| GPi001 | L-12 | 94.50 | 98.50 | 90.50 | 94.90 | 79.00 | 78.00 | 80.00 | 78.61 |
| GPi071 | L-12 | 90.75 | 88.50 | 93.00 | 90.49 | 75.00 | 78.00 | 72.00 | 75.50 |
| GPi108 | R-12 | 89.00 | 91.00 | 87.00 | 89.37 | 81.25 | 82.50 | 80.00 | 81.55 |
| GPi153 | R-12 | 88.25 | 94.00 | 82.50 | 88.93 | 82.75 | 80.50 | 85.00 | 81.57 |
| **Avg** | | 90.13 | 90.71 | 89.56 | 90.13 | 79.65 | 79.32 | 79.97 | 79.29 |
| **Std** | | 2.35 | 4.22 | 3.78 | 2.47 | 3.31 | 3.77 | 3.99 | 3.36 |

*L-12* Left channel-12, *L-23* Left channel-23, *L-34* Left channel-34, *R-12* Right channel-12, *R-23* Right channel-23, *R-34* Right channel-34. *Avg* Average. *Std* Standard deviation. *ACC* accuracy, *Sen* sensitivity, *Spe* specificity, *F1* F1-score.

| **Supplementary Table 9** Subject-wise statistical comparison of electrophysiological features between spindle and non-spindle events within the LFP channels with the highest R-map match. | | | |
| --- | --- | --- | --- |
| **Patients** | **Channel Selected** | **Statistics of electrophysiological signals** | |
|  |  | **Power** | **PLV** |
| *STN-Lead* | | | |
| STN004 | L-12 | U = 26841, p = 1.29×10^-32^ | U = 24589, p = 1.52×10^-21^ |
| STN-010 | L-12 | U = 3641, p = 4.89×10^-6^ | U = 3689, p = 1.88×10^-6^ |
| STN-011 | R-34 | U = 947, p = 0.020 | U = 1025, p = 0.002 |
| STN-063 | R-34 | U = 4390, p = 3.15×10^-8^ | U = 4363, p = 5.53×10^-8^ |
| STN-070 | R-12 | U = 5141, p = 4.65×10^-10^ | U = 5306, p = 1.18×10^-11^ |
| STN-084 | L-12 | U = 16564, p = 2.40×10^-26^ | U = 13991, p = 1.59×10^-11^ |
| STN-090 | L-12 | U = 19944, p = 3.04×10^-7^ | U = 21915, p = 5.14×10^-13^ |
| STN-112 | L-34 | U = 726, p = 0.004 | U = 738, p = 0.002 |
| STN-116 | L-12 | U = 4664, p = 0.008 | U = 5172, p = 2.98×10^-5^ |
| STN-117 | L-12 | U = 5137, p = 2.04×10^-6^ | U = 5398, p = 2.65×10^-8^ |
| STN-143 | L-12 | U = 1009, p = 0.468 | U = 1013, p = 0.447 |
| STN-150 | L-12 | U = 20660, p = 1.35×10^-22^ | U = 18818, p = 4.89×10^-14^ |
| STN-171 | R-12 | U = 5035, p = 1.76×10^-6^ | U = 4737, p = 1.26×10^-4^ |
| *GPi-Lead* | | | |
| GPi-001 | L-34 | U = 4755, p = 1.27×10^-7^ | U = 4881, p = 9.76×10^-9^ |
| GPi-071 | L-34 | U = 42161, p = 7.11×10^-28^ | U = 36813, p = 2.70×10^-15^ |
| GPi-108 | L-34 | U = 2190, p = 5.19×10^-8^ | U = 2138, p = 3.28×10^-7^ |
| GPi-153 | R-34 | U = 2063, p = 9.09×10^-10^ | U = 1832, p = 7.35×10^-6^ |

*L-12* Left channel-12, *L-23* Left channel-23, *L-34* Left channel-34, *R-12* Right channel-12, *R-23* Right channel-23, *R-34* Right channel-34. *PLV* Phase-locking value. *U S*tatistics of Mann-Whitney U rank test.

| **Supplementary Table 10** Statistical analysis of paired VTA overlap differences across STN subregions | | | | | | | | | |
| --- | --- | --- | --- | --- | --- | --- | --- | --- | --- |
| Subregions | Motor | | | Associative | | | Limbic | | |
|  | ΔVolume | W-stat | *P* value | ΔVolume | W-stat | *P* value | ΔVolume | W-stat | *P* value |
| 1.0 mm | -0.0885 | 40.00 | 0.9697 | -0.5725 | 10.00 | 0.0840 | 0.3727 | 30.00 | 0.1094 |
| 1.5 mm | -0.1859 | 46.00 | 1.0000 | **-1.9781** | **7.00** | **0.0371** | 1.2892 | 30.00 | 0.1094 |
| 2.0 mm | -0.5398 | 46.00 | 1.0000 | **-4.3452** | **16.00** | **0.0398** | **2.8479** | **78.00** | **0.0215** |
| 2.5 mm | -1.6775 | 42.00 | 0.8394 | **-7.1039** | **16.00** | **0.0398** | **4.7408** | **77.00** | **0.0266** |
| 3.0 mm | -2.9814 | 40.00 | 0.7354 | **-9.1065** | **17.00** | **0.0479** | **6.7566** | **77.00** | **0.0266** |
| 3.5 mm | -4.0217 | 37.00 | 0.5879 | -10.0804 | 18.00 | 0.0574 | **8.6183** | **77.00** | **0.0266** |
| 4.0 mm | -5.0447 | 36.50 | 0.5540 | -10.1959 | 22.00 | 0.1099 | **9.7552** | **76.00** | **0.0327** |
| 4.5 mm | -5.7671 | 35.00 | 0.4973 | -9.7962 | 25.00 | 0.1677 | 10.4113 | 72.00 | 0.0681 |
| 5.0 mm | -6.3315 | 35.00 | 0.4973 | -8.5675 | 26.00 | 0.1909 | 10.2770 | 66.00 | 0.1677 |
| 5.5 mm | -6.5526 | 35.00 | 0.4973 | -6.8540 | 26.00 | 0.1909 | 9.0893 | 63.00 | 0.2439 |
| 6.0 mm | -6.3765 | 34.00 | 0.4548 | -4.8833 | 29.00 | 0.2734 | 7.0957 | 63.00 | 0.2439 |
| 6.5 mm | -5.8204 | 25.00 | 0.5195 | -2.8586 | 31.00 | 0.3396 | 5.1225 | 57.00 | 0.4548 |
| 7.0 mm | -5.1487 | 14.00 | 0.3594 | -1.3629 | 34.00 | 0.4548 | 3.7940 | 56.00 | 0.4973 |
| 7.5 mm | -4.4918 | 8.00 | 0.3750 | -0.6127 | 31.00 | 0.8984 | 2.7021 | 57.00 | 0.4548 |
| 8.0 mm | -3.5269 | 9.00 | 0.4688 | -0.3293 | 28.00 | 1.0000 | 1.5513 | 59.00 | 0.3757 |
| 8.5 mm | -2.4990 | 4.00 | 0.4375 | -0.1884 | 9.00 | 0.8438 | 0.6897 | 33.00 | 0.6250 |
| 9.0 mm | -1.6857 | 1.00 | 0.2500 | -0.1016 | 3.00 | 1.0000 | 0.1646 | 35.00 | 0.4922 |
| 9.5 mm | -1.0353 | 1.00 | 0.2500 | -0.0287 | 1.00 | 1.0000 | -0.1253 | 28.00 | 0.5703 |
| 10.0 mm | -0.5529 | 0.00 | 0.5000 | 0.0000 | 0.00 | 1.0000 | -0.0876 | 9.00 | 0.8125 |

*W-stat* Non-parametric paired Wilcoxon signed-rank test. Significant results (P < 0.05) are highlighted in bold.

| **Supplementary Table 11** Predictive decoding performanceof MetaCNN across varying temporal horizons | | | | |
| --- | --- | --- | --- | --- |
| Temporal window (Δ*t*) | **ACC** | **Sen** | **Spe** | **F1** |
| 0.0 s | 83.51 ± 3.80 | 83.42 ± 5.44 | 83.47 ± 5.83 | 83.30 ± 3.13 |
| -0.5 s | 75.60 ± 4.51 | 69.15 ± 4.97 | 64.24 ± 8.00 | 67.38 ± 4.54 |
| -1.0 s | 68.71 ± 5.50 | 69.65 ± 5.76 | 67.18 ± 8.87 | 68.73 ± 5.56 |
| -1.5 s | 68.59 ± 5.44 | 68.74 ± 5.43 | 68.44 ± 8.44 | 68.16 ± 5.48 |
| -2.0 s | 64.87 ± 5.59 | 73.21 ± 6.27 | 67.12 ± 6.94 | 70.84 ± 5.13 |

Data are presented as Mean ± Standard Deviation across 17 test subjects. The observation window corresponds to a 2-second local field potential (LFP) segment systematically shifted backward relative to the true spindle onset (Δ*t* = 0.0 s).

| **Supplementary Table 12** Symmetric matrix of pairwise statistical comparisons for decoding accuracy across temporal horizons. | | | | | |
| --- | --- | --- | --- | --- | --- |
| Δ*t* | 0.0 s | -0.5 s | -1.0 s | -1.5 s | -2.0 s |
| 0.0 s | - | 3.58 × 10^-4^ | 4.74 × 10^-6^ | 1.64 × 10^-6^ | 1.17 × 10^-8^ |
| -0.5 s | 3.58 × 10^-4^ | - | 0.0042 | 0.0218 | 3.47 × 10^-4^ |
| -1.0 s | 4.74 × 10^-6^ | 0.0042 | - | 1.0000 | 0.8398 |
| -1.5 s | 1.64 × 10^-6^ | 0.0218 | 1.0000 | - | 0.6301 |
| -2.0 s | 1.17 × 10^-8^ | 3.47 × 10^-4^ | 0.8398 | 0.6301 | - |

Values represent Bonferroni-corrected *P*-values derived from pairwise paired *t*-tests comparing decoding accuracy between different temporal shifts. The symmetric matrix format illustrates the significance of accuracy degradation as the prediction horizon extends. Significance levels: * *P* < 0.05, ** *P* < 0.01, *** *P* < 0.001.
